# The impact of introducing meningococcal C/ACWY booster vaccination among adolescents in Germany: a dynamic transmission modelling study

**DOI:** 10.1101/2024.12.19.24319393

**Authors:** Felix Günther, Ulrich Reinacher, Sarah Chisholm, Matas Griskaitis, Michael Höhle, Stefan Scholz, Viktoria Schönfeld, Ole Wichmann, Thomas Harder, Frank G. Sandmann

**Author notes:** Corresponding author: Felix Günther, Immunization Unit - Team Vaccine/VPD Modelling Department for Infectious Disease Epidemiology Robert Koch Institute, Nordufer 20, 13353 Berlin Germany.

## Abstract

**Background:** In Germany, primary vaccination against invasive meningococcal disease (IMD) serogroup C aims to reduce the highest burden of IMD in infants aged 12-23 month. Due to another IMD-peak in adolescents, we modelled the potential impact of introducing adolescent boosters with conjugate meningococcal C or ACWY (MenC/MenACWY) vaccines.

**Methods:** We built an age- and serogroup-structured dynamic-transmission model for Germany, which we calibrated to national surveillance data in 2005-2019. We simulated five vaccination scenarios of either continuing with the current MenC primary vaccination (scenario 1), or additionally introducing MenC or MenACWY boosters at age 13 years (scenarios 2-3) or 16 years (scenarios 4-5). We performed comprehensive sensitivity analyses, including on the protection against carriage and serotype replacement.

**Results:** The calibrated model projected for scenario 1 an annual mean of 243 (95%-uncertainty interval: 220-258) expected IMD cases over a 10-year period. Introducing the MenC booster prevented an estimated 5 (3.9-6.7) and the MenACWY booster 8 (6.7-9.1) IMD cases per year on average (scenario 2 and 3). The number-needed-to-vaccinate (NNVs) to prevent one IMD case were 140,000 (100,000-180,000) and 91,000 (76,000-100,000), respectively. To prevent one sequela or death, NNVs were higher (i.e., less efficient). Results were broadly similar for scenarios 4-5. Simulations suggested relevant serotype replacement starting eight-to-ten years after introducing the MenACWY booster.

**Conclusions:** Introducing adolescent MenC or MenACWY boosters marginally reduces the expected IMD burden in Germany. Effectiveness and efficiency of evaluated strategies depend on future incidence. The magnitude of future serotype replacement for the MenACWY vaccine is highly uncertain.

## Introduction

Invasive meningococcal disease (IMD) can cause serious bacterial meningitides and septicaemia, with fatal outcome in about 10% (1) and long-term health consequences in up to 20% of cases (2). IMD is caused by the gram-negative bacteria meningococcus or *Neisseria meningitidis*, which can be transmitted through droplets from carriers mixing with susceptible individuals. Meningococcus carriage is widespread in the general population, with an estimated carriage prevalence of about 10% in healthy individuals and pronounced variation by age (3).

In Germany, national surveillance data showed an annual incidence of 0.4 IMD cases per 100,000 people between 2012-2015 (1). The highest proportion of cases was seen in children aged less than 5 years (29.9% of all cases, particularly in the first two years of life), with a second peak in the incidence of IMD cases observed in adolescents aged 15-19 years (14.3% of all cases) (1). Of the 12 serogroups in total, the IMD cases in Germany were most frequently caused by serogroup B (MenB) with an annual incidence of 0.27/100,000 population, followed by MenC (0.08/100,000), MenW (0.02/100,000) and MenY (0.03/100,000). The overall case-fatality was 9.6% across serogroups, ranging from 9.4% for MenB to 13.6% for MenC (1). Similar to other countries, an increase has been observed in IMD case numbers caused by MenW (all ages) and MenY (primarily in adolescents) (4). The incidence of MenC cases has been decreasing continuously ever since 2007 following the introduction of routine MenC vaccination (1, 4).

Primary vaccination against IMD serogroup C in infants aged 12-23 month is recommended in Germany since 2006 (5). The MenC vaccine coverage in school-aged children increased from 53% in 2008 (6) to 90 % in 2020 (7). Data suggest that the level of protection following primary MenC vaccination declines over time (8, 9), which may contribute to the second peak in IMD cases observed in adolescents (10). As a consequence, for example the UK recommends adolescent MenC booster vaccination against MenC since 2014 and -in response to a rise in MenW cases-against MenACWY since 2016(11).

This study quantified the potential impact of introducing an adolescent booster program with conjugate meningococcal monovalent (MenC) or polyvalent (MenACWY) vaccines in the national immunization schedule of Germany. The results aimed to inform the German Standing Committee on Vaccination (STIKO), which is the national immunization technical advisory group (NITAG) in Germany.

## Methods

The key methodological aspects of this modelling study are summarized below, more details can be found in the Supplementary texts S1 and S2.

### Dynamic-transmission model of meningococcal carriage

We developed a time-continuous, deterministic Susceptible-Infected-Susceptible (SIS)-type dynamic-transmission model that was structured into 86 age groups (0, 1, 2,…, 84, and 85+ years). Ageing and vaccination coverage are implemented as discrete, annual step changes. Vaccination provides (partial) protection against becoming a carrier (via a reduction of the force of infection) and/or protection against IMD among new carriers and wanes over time. Expected yearly IMD cases by age- and serogroup are derived from the model by scaling incident new carriers with age- and serogroup-specific case-carrier ratios. The design and structure of the model for Germany was informed by a similar model developed for the UK (12).

At any time, individuals of all ages are in one of 20 different compartments that represent four groups of carriage status (i.e., susceptible, MenC carrier, MenAWY carrier or ‘MenB/Other’ carrier), three groups of vaccination status (unvaccinated, vaccinated after primary or booster vaccination, and waned vaccine-derived protection) and two meningococcal conjugate vaccines (monovalent MenC or polyvalent MenACWY); see Figure 1. Demographic turnover was informed by population statistics and prospective projections, with new-borns allocated to the unvaccinated and susceptible compartment of age 0 each year. For more details on the mathematical model see SuppText S1.

**Figure 1:**
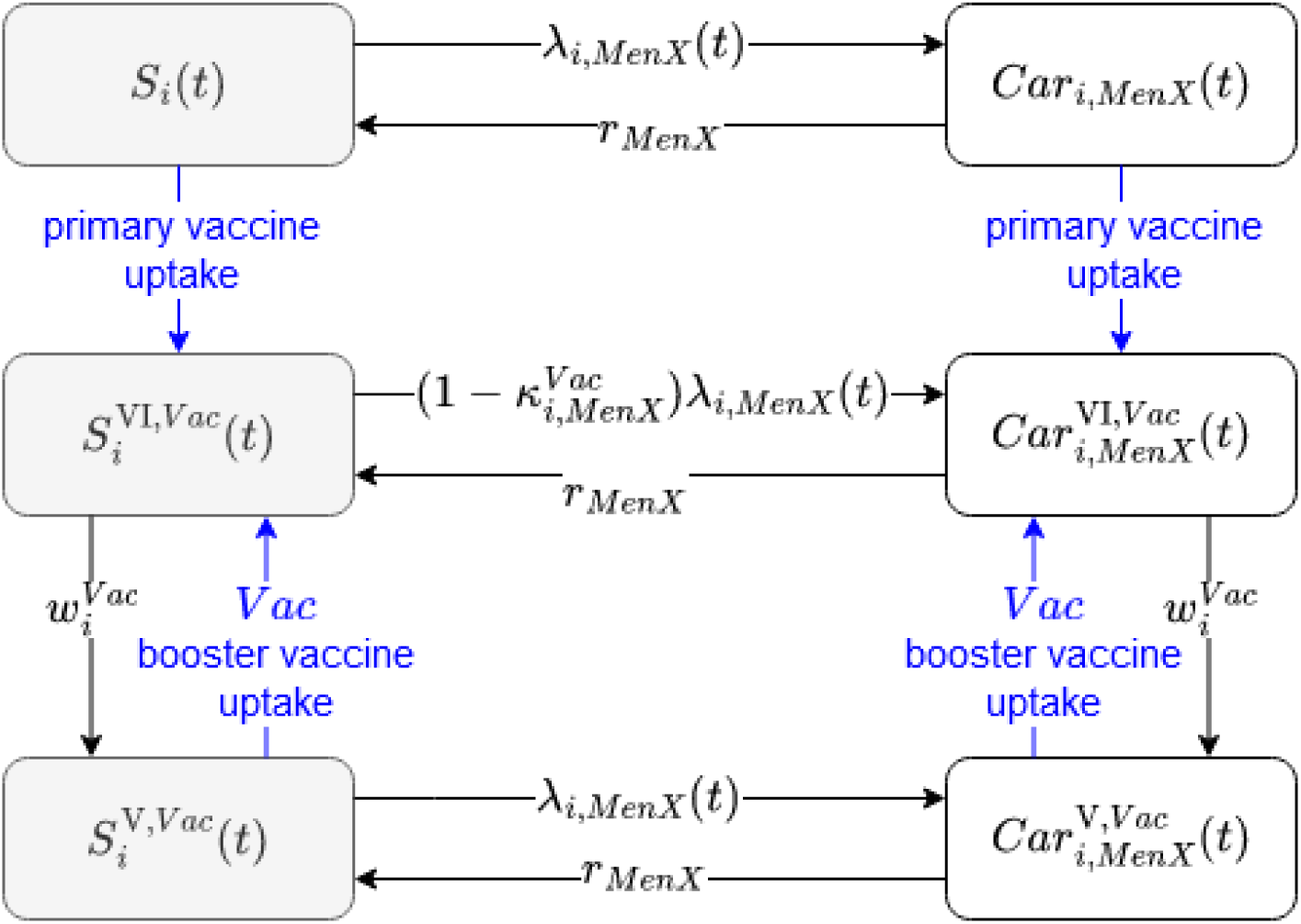
Diagram of the dynamic-transmission model of meningococcal carriage in Germany. The model tracks 86 age groups (denoted with i), three groups of serogroups (denoted with ‘MenX’ that represent MenC, MenAWY or MenB/Others) and two vaccines (denoted with ‘Vac’ that can represent MenC or MenACWY vaccines). Black arrows indicate continuous changes in the model, blue arrows indicate discrete changes at specific time points. Individuals in the susceptible compartments can become carriers based on the time-dependent force of infection by age and serogroup, denoted by λ. In the carriage compartments, individuals can recover to become fully susceptible again. After primary vaccination the vaccinated non-carriers are (partially) protected against becoming a newly-infected carrier by reducing the force of infection based on a factor κ, specific to a certain age, vaccine type, and serogroup. For incident carriers, vaccination can additionally protect against IMD by reducing the case-carrier ratio (not shown in diagram). Given the waning of immunity individuals lose the vaccine-derived protection exponentially over time based on rate ω, specific to age and vaccine type. These individuals can become protected again through a booster vaccine. Demographic turnover was informed by population statistics (historically) and projections (prospectively).

### Model input data and calibration

We parametrized the model by utilizing published data from literature as well as a calibrating the model to German surveillance data. Protection among vaccinated individuals was mainly parametrized based on vaccine effectiveness estimates from post-licensure surveillance in UK, the case-carrier ratios were specified using a model-based comparison of published estimates of meningococcal carriage prevalence and IMD case numbers from Germany. Age-related contact rates, mean carriage duration, and estimates on the probability of sequelae and death among IMD cases were extracted from external data. An overview of input data is given in SuppTab. 1, more details in SuppText S2.

Conditional on these parameters and assumptions, we estimated parameters governing the sero- and age group specific force of infection during model calibration. For this, we fitted the model to age- and serogroup specific yearly IMD case numbers in Germany from 2005-2019 using a negative binomial-based maximum likelihood estimation. We explored four model-specifications with varying complexity and performed model selection against Akaike and Bayesian information criteria (AIC, BIC). More details on the calibration are given in SuppText S1.

### Simulating future vaccination scenarios in Germany over 10 years, 2020-2029

After calibrating the model, we simulated five future vaccination scenarios over 10 years:

- Scenario 1: Primary vaccination with MenC in the second year of life (month 12-23), i.e. status quo
- Scenario 2: Scenario 1 + MenC booster vaccination at age 13 years
- Scenario 3: Scenario 1 + MenACWY booster vaccination at age 13 years
- Scenario 4: Scenario 1 + MenC booster vaccination at age 16 years
- Scenario 5: Scenario 1 + MenACWY booster vaccination at age 16 years

All simulations started in 2020 with the model compartments initialized based on the calibrated model at the end of the calibration period 2019. Scenario 1 follows the current vaccination recommendation in Germany, with observed vaccine coverage of 80%. In the main analysis we assumed all individuals who received the primary vaccination to receive the booster, which we lowered in sensitivity analysis (see below).

### Estimating the effectiveness and efficiency of introducing the adolescent booster

We quantified results by age and serogroup in terms of the number of expected IMD cases with and without long-term health consequences (sequela) as well as deaths. Probabilities for 16 different sequelae were sourced from the published literature (13, 14). For deaths we multiplied cases with the estimated case-fatality ratios (CFRs) specific to age and serogroup in Germany (SuppText S1).

Afterwards, we estimated the associated number-needed-to-vaccinate (NNVs) to prevent one case with or without sequelae or death by comparing the results from the booster vaccination (scenarios 2-5) with the results of the no-booster baseline (scenario 1, status quo). Our results also show an uncertainty interval, which we derived from the parameter uncertainty of the dynamic-transmission model by sampling 100 sets of parameters from the approximate multivariate normal distribution of the maximum likelihood estimate (obtained as a result from the calibration) and then solving the model for each of the 100 parameter combinations. The uncertainty was propagated throughout the modelling to express a 95%-uncertainty interval (95%-UI) for quantities of interest.

### Sensitivity analysis of key parameters and methodological choices

We explored the impact of key parameters and methodological choices in sensitivity analyses. First, we increased/decreased the estimated transmission parameters by −/+ 20% (sensitivity A1 and A2). We also explored keeping the future IMD incidence constant at pre-2020 levels before applying the relative reductions obtained from the dynamic model in the main analysis (A3) to minimize the effect of the downward-trend already observed for MenC in Germany until 2019. Second, a structural analysis simulated results based on second-best-fitting model (B1). Third, due to substantial uncertainty regarding the VE against meningococcal carriage for the MenACWY vaccine, we varied the protection against carriage from 0 to 80% (C). Fourth, we explored further parameter uncertainties by (i) reducing the mean duration of protection after the booster vaccination from 10 to 4 years (D1), (ii) increasing the mean duration of carriage from 6 (15) to 12 months (16) (D2), and using two alternative specifications for the CCRs and corresponding initialization of the model (D3 and D4). Fifth, we investigated the effect of a reduced booster uptake of 60% among all primary vaccinated individuals, leading to an overall uptake around 50% among adolescents, roughly corresponding to the complete HPV vaccine uptake among female adolescents in Germany (E). Lastly, we explored separately a longer time horizon of 30 years to capture the serotype replacement that only started to occur towards the end of the 10-year simulation period.

All analyses were performed in R based on an analysis pipeline implemented using the *targets* package (17). The transmission model was implemented using the function *lsoda* from the *deSolve* package (18) Model code will be made public upon final publication of the manuscript.

## Results

### Model calibration to historical surveillance data

The best-fitting model used age- and serogroup-specific transmission parameters for eight age subgroups (model 2b; SuppTab. 2 for model selection and parameter estimates).

Compared to the observed average number of 371 IMD cases per year in 2005-2019, our model predicted 366 (95%-UI: 346-388) IMD cases annually during the calibration period. The estimated number of IMD cases fit the observed trends in all serogroups (Fig. 2A), replicating the observed decline in serogroup C and B/Other, and the increase in serogroup AWY in 2016-2019. The model also captured trends of IMD cases in age subgroups well (Fig. 2B; SuppFig. 1). The calibrated model reflected introduction of the MenC primary vaccination after 2005; e.g., about 80% of individuals born in 2010 were vaccinated by 2013 and the meningococcal carriage prevalence was low (Fig. 2C). Individuals born in 2000 were older at the start of the primary vaccination program, had a lower vaccine uptake and showed a clear increase in carriage after age 15. Carriage prevalence by age changed during the calibration period with increasing prevalence for MenACWY and a decrease for serogroup Other/B (SuppFig. 2).

**Figure 2:**
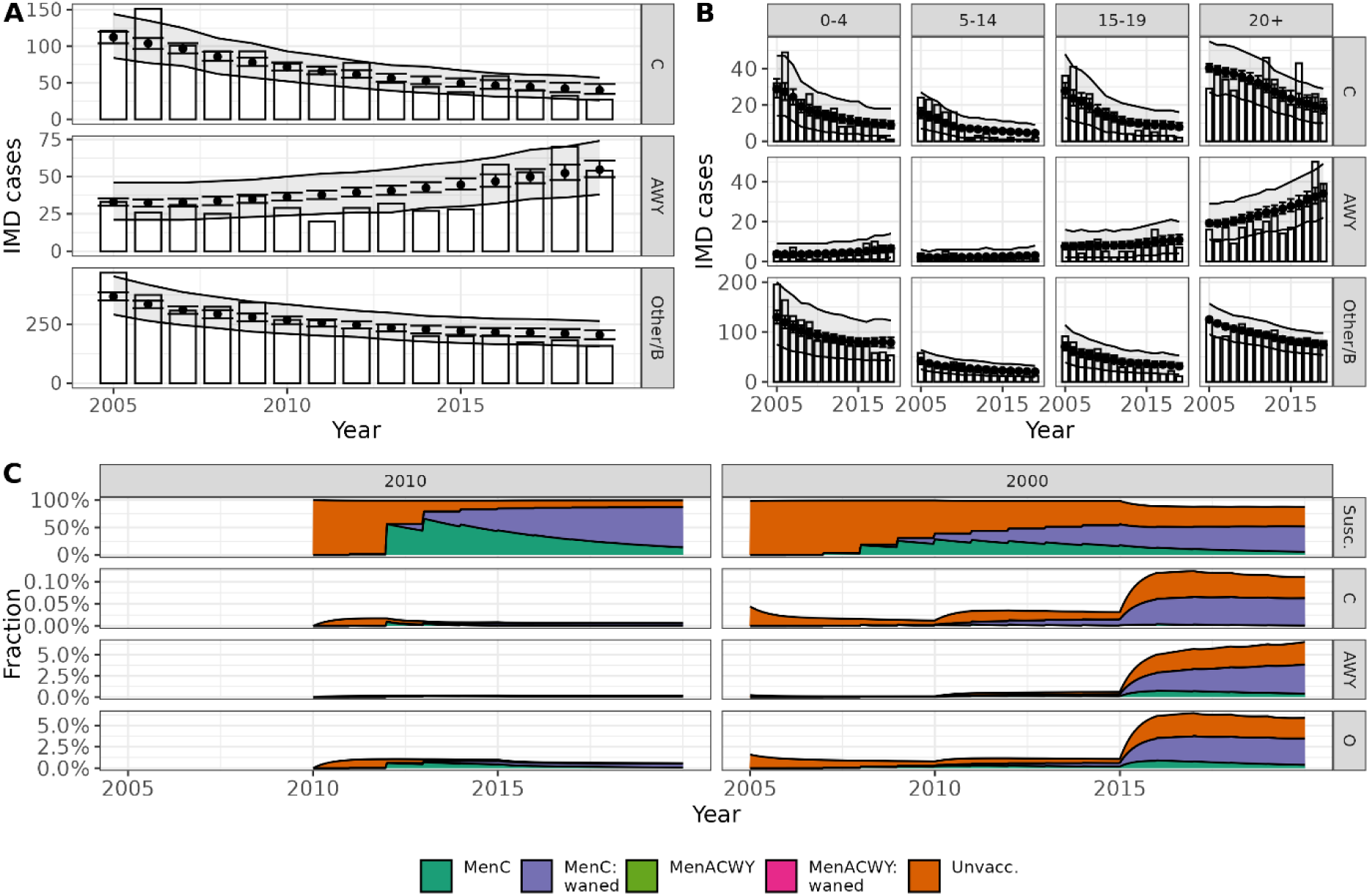
Calibration results of the model fit to the national surveillance data in Germany in 2005-2019. Panel A and B: Annual expected number with 95% uncertainty interval of IMD cases per serogroup (dots and error bars), 95%-uncertainty interval based on the fitted negative binomial distribution (light-grey ribbon), and observed IMD cases (bars), shown for the total population (panel A) and separated in 4 age groups (panel B). Panel C: Relative proportion of all individuals in two exemplary birth cohort born in 2010, 2000 (per column) in the compartments of susceptible individuals and carriers per serogroup (per row) and according to vaccination status reflecting the historic routine data (colour coding). Vaccination was (only) recommended with the MenC vaccine during the calibration period. Note the different y-axes limits.

### Simulation of adolescent booster vaccination strategies

Aggregated over the 10-year simulation period and all age groups, MenC booster vaccination at age 13 (scenario 2) reduced the expected number of IMD cases with serogroup C from 289 to 238 by 18% (95%-UI: −20%, −16%), with minimal changes in the other serogroups (Fig 3A). The MenACWY booster (scenario 3) reduced MenC-IMD cases from 289 to 265 (−8%, 95%-UI: −10%, −7%), and also MenAWY-IMD cases from 655 to 589 (−10%, 95%-UI: −12%, −9%). However, for serogroup Other/B, simulations indicated a small increase in Other/B IMD cases from 1481 to 1494 by 1% (95%-UI: +0.8%, +1.2%). This serotype replacement in the MenACWY booster scenario became apparent towards the end of the simulation period (Fig. 3B-3C). We observed not only direct but also indirect effects among age-groups not directly targeted by the adolescent booster (Fig. 3C). The scenarios of older adolescent age (booster at age 16, scenarios 4 and 5) indicated slightly larger direct and indirect effects compared to the booster at age 13 (Fig. 3A).

**Figure 3:**
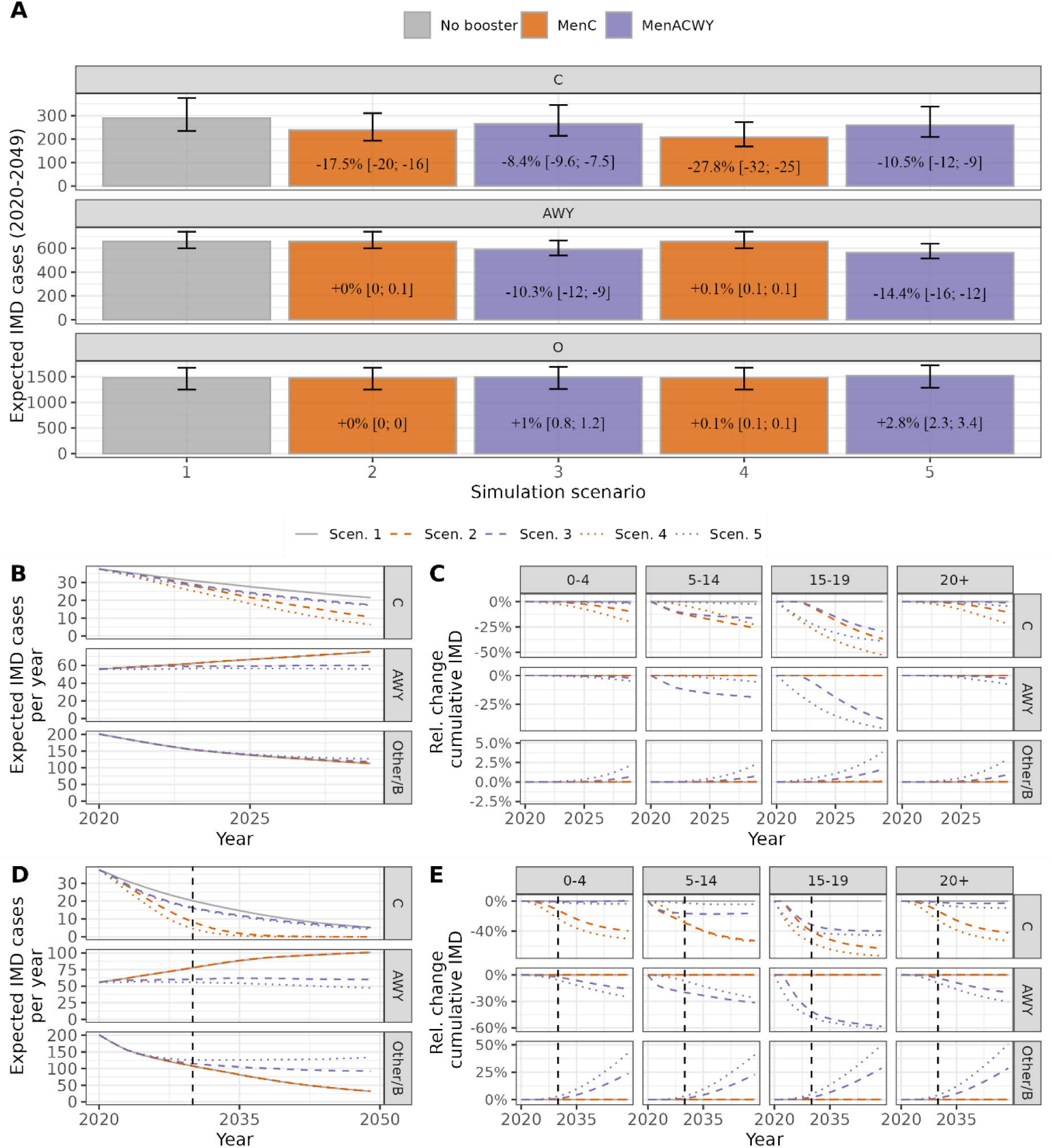
Expected number of IMD cases by serogroup in each vaccination scenario 1-5 over 10 years, 2020-2029. Panel A: Cumulative total number of cases and relative change compared to scenario 1. Panel B: annual number of expected cases by serogroup. Panel C: relative change in the cumulative number of cases by sero- and age group. Panel D+E: same as B+C but with 30-year simulation period. *During the simulation period, trends from the calibration period were continued: the general downward trend of expected MenC-IMD cases was accelerated by the introduction of an adolescent booster, more pronounced for MenC than for MenACWY booster. Without or with the MenC booster, a further increase in AWY-IMD cases was simulated, whereas with the MenACWY booster, expected annual AWY-IMD numbers remained relatively constant during the simulation period. Other/B-IMD cases were decreasing in all scenarios, with the MenACWY booster, there was, however, an increase in expected cases compared to the no booster/MenC booster scenario that started towards the end of the 10-year simulation period (“serotype replacement”)*.

For the MenC booster, we simulated considerable reductions in MenC-carriage among adolescents directly targeted by the booster strategies (e.g., birth cohort 2010) compared to no booster (Fig. 4A, Scen. 2 vs. Scen. 1). For the MenACWY booster, effects on carriage were less pronounced (Fig. 4A, Scen. 3). Individuals from the 2000 birth cohort were not targeted by the booster vaccination starting in 2020. However, at the carrier level, slight indirect effects can be seen towards the end of the simulation period (reduction in MenC carriage with MenC booster; slight reduction in MenC and ACWY, and small increase in Other/B carriage with the MenACWY booster, Fig. 4B). Note that carriage prevalence of the MenAWY serogroup was substantially higher than for MenC (SuppFig. 3). This explains larger replacement effects with the MenACWY booster.

**Figure 4:**
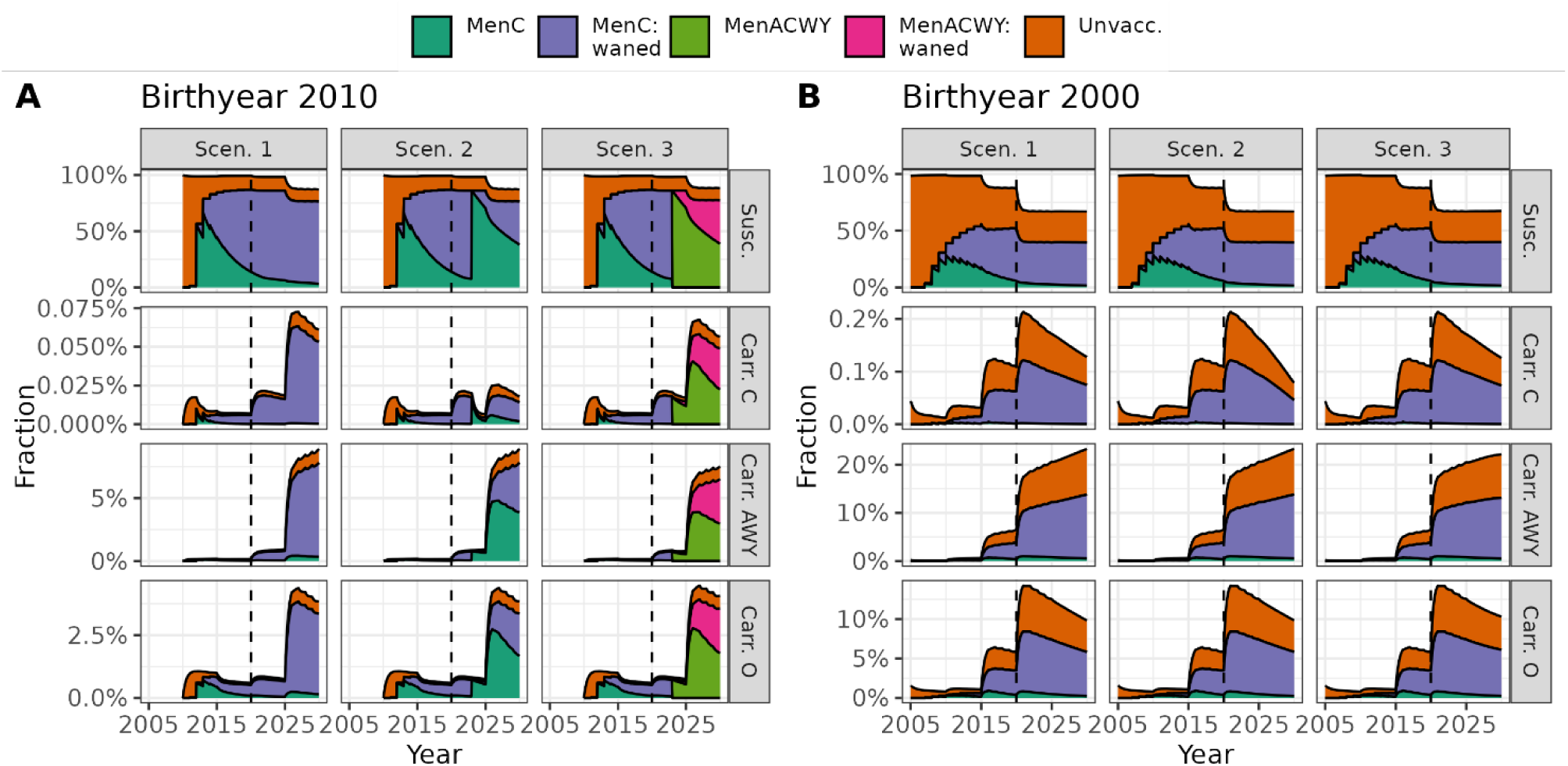
Illustration of simulation results on carriage prevalence level for vaccination scenario 1-3 for two selected birth cohorts over 10 years, 2020-2029. Relative proportion of all individuals in two exemplary birth cohorts born in 2010 (panel A), and 2000 (panel B) for vaccination scenarios 1-3 (columns) in the compartments of susceptible individuals and meningococcal carriage status (per row) and according to vaccination status based on the simulation assumptions (colour coding). Dotted vertical line marks change from calibration to simulation period in year 2020. Note the different y-axis scales in each row and panel.

### Effectiveness and efficiency of introducing the adolescent booster

There was a total of 2425 (95%-UI: 2199-2580) expected IMD cases, 987 (896–1048) cases with sequelae, and 219 (199–233) deaths during the 10-year period in scenario 1. On average, the adolescent MenC booster (scenario 2) prevented 5.0 expected IMD cases (3.9-6.7), 2.1 cases with sequalae (1.7-2.8) and 0.6 deaths (0.5-0.8) annually over the 10 years (Table 1). The adolescent MenACWY booster (scenario 3) prevented an estimated total of 7.7 IMD cases (6.7-9.1), 3.1 IMD cases with sequalae (2.7-3.7) and 0.7 deaths (0.6-0.8) annually over 10 years. The respective number-needed-to-vaccinate (NNV) to prevent one case were 140,000 (100,000–180,000) for scenario 2 and 91,000 (76,000–100,000) for scenario 3. To prevent one sequela or death the NNVs were larger (Table 1). The results for scenarios 4 and 5 indicated that a booster vaccination at older age (at age 16 instead of 13) can be slightly more effective and efficient than scenarios 2 and 3 (Table 1), but uncertainty intervals were overlapping. For serogroup-specific results and overall efficacy focusing on serogroups C and AWY see SuppTab. 3 and 4.

**Table 1:** Effectiveness and efficiency of introducing the adolescent booster vaccination program in Germany over 10 years (as compared to no-booster program, scenario 1).

| Scenario | Outcome | Prevented total number of outcomes (Est. + 95%-UI) |  |  | Numbers needed to vaccinate to prevent one outcome (Est. 95%-UI) |  |  |
| --- | --- | --- | --- | --- | --- | --- | --- |
|  |  | Est. | Lower | Upper | Est. | Lower | Upper |
| 2 | IMD cases | 50 | 39 | 67 | 140,000 | 100,000 | 180,000 |
|  | Sequalae or death | 21 | 17 | 28 | 330,000 | 250,000 | 420,000 |
|  | Death | 6 | 5 | 8 | 1,200,000 | 890,000 | 1,500,000 |
| 3 | IMD cases | 77 | 67 | 91 | 91,000 | 76,000 | 100,000 |
|  | Sequalae or death | 31 | 27 | 37 | 220,000 | 190,000 | 260,000 |
|  | Death | 7 | 6 | 8 | 1,000,000 | 840,000 | 1,200,000 |
| 4 | IMD cases | 80 | 61 | 100 | 84,000 | 64,000 | 110,000 |
|  | Sequalae or death | 34 | 26 | 44 | 200,000 | 150,000 | 260,000 |
|  | Death | 10 | 7 | 13 | 700,000 | 530,000 | 910,000 |
| 5 | IMD cases | 83 | 71 | 100 | 80,000 | 67,000 | 93,000 |
|  | Sequalae or death | 34 | 29 | 41 | 190,000 | 160,000 | 230,000 |
|  | Death | 8 | 7 | 10 | 810,000 | 680,000 | 950,000 |
| IMD: invasive meningococcal disease, NNV: number needed to vaccinate, Est.: point estimate. UI: uncertainty interval.<br>Scenario 2: MenC booster at age 13; Scenario 3: MenACWY booster at age 13; Scenario 4: MenC booster at age 16; Scenario 5: MenACWY booster at age 16.<br>The prevented total number of outcomes is the difference in the expected cumulative numbers with scenarios 2-5 as compared to scenario 1 (i.e., no-booster vaccination in adolescents) over 10 year, 2020-2029. The outcome “sequalae or death” considered at least 1 sequalae or fatal progression. The NNV (last column) was calculated based on an additional 6.963 million administered booster vaccine doses in scenarios 2 and 3 (vaccination at age 13), and an additional 6.671 million booster doses in scenarios 4 and 5 (age 16). |  |  |  |  |  |  |  |

### Sensitivity analyses of simulation results

The sensitivity analyses of key assumptions revealed that simulation results were sensitive to the transmission dynamics assumed during the simulation period, with smaller (serotype-specific) transmission and expected IMD case counts reducing effectiveness and efficiency of the booster vaccinations, while higher expected case counts led to higher effectiveness and efficiency (sensitivity analyses A, Fig. 5, SuppFig. 4). While the overall expected IMD case numbers without booster vaccination remained relatively stable in the sensitivity analyses B-E, changes in (structural) assumptions had (sometimes complex) consequences for the simulated effects of the booster vaccination with different implications for the MenC and MenACWY booster (Fig. 5, SuppFig. 4). Particularly relevant with respect to structural uncertainty were assumptions on the CCRs (e.g., analysis D4) and - for the MenACWY booster – the degree of protection against carriage, as these assumptions directly affected the magnitude and type of indirect effects. Reducing the booster uptake to 60% among all individuals with primary immunization reduced the expected number of prevented cases. As the number of vaccine doses was also reduced, efficiency of the booster vaccination programs remained relatively unchanged. A more detailed description of the results is given in SuppText S3, simulated effects of the booster vaccinations in each sensitivity analysis are visualised in SuppFigs. 5-17.

**Figure 5:**
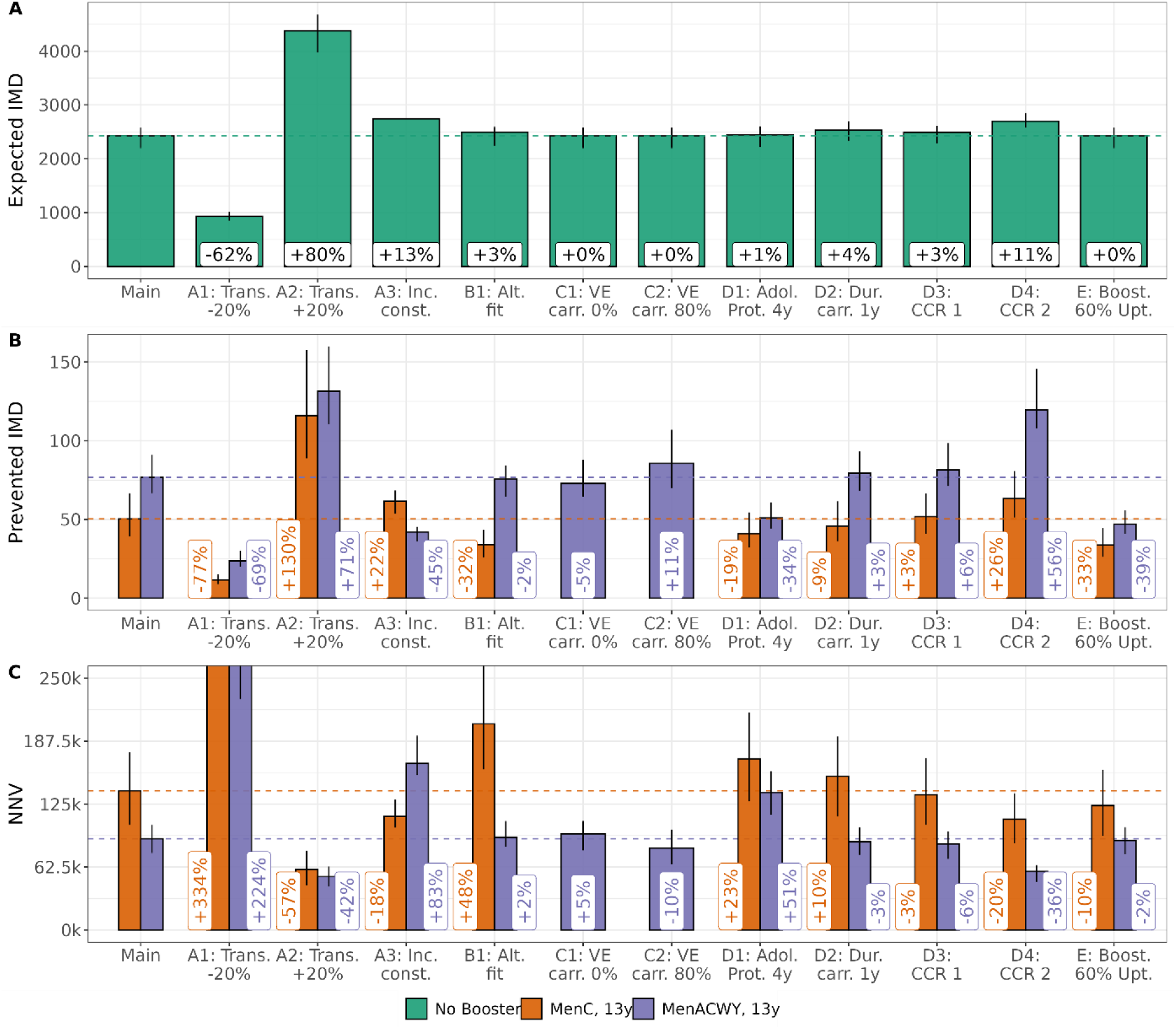
Results of the sensitivity analyses for booster vaccination at age 13. Panel A: Expected IMD cases during the simulation period without booster vaccination for each (sensitivity) analysis and relative change compared to main analysis. Panel B: reduction of expected IMD cases based on the booster vaccination program at age 13 (C or ACWY vaccine, colour coded) for each (sensitivity) analysis and relative change compared to prevented cases in the main analysis. Panel C: corresponding NNVs and relative change compared to main analysis. Dotted lines show results in main analysis for visual comparison.

### Extended simulation period of 30 years

To analyse possible long-term effects of the introduction of adolescent booster vaccines, we have increased the simulation period from 10 to 30 years in additional analyses. For the MenC booster, results were comparable to the 10-year simulation, with a slight improvement in efficiency due to longer accumulation of indirect effects (e.g., mean number of yearly prevented expected IMD cases based on the booster at age 16: 9.7 (6.7, 14.7) vs. 8.0 (6.1, 10.0) in the 10-year period; NNV: 69,000 (45,000, 100,000) vs. 84,000 (64,000, 110,000); Fig 3D+E, SuppTab. 5, SuppFig. 19). For the MenACWY booster, the results differed more markedly between the 10- and 30-year simulations. Aggregated over a 30-year period, the serotype replacement of MenAWY by serogroup Other/B, which only manifested itself sometime after the introduction of the booster vaccination, played a markedly greater role (Fig 3D+E). The prevented AWY cases were largely replaced by additional expected Other/B cases; for the ACWY booster in year 16, the expected overall effect of introducing the booster vaccination was even negative (SuppTab. 5 and 6, SuppFig. 18).

The magnitude of serotype-replacement, and consequently the efficiency of the MenACWY booster in terms of prevented IMD cases over the 30-year simulation period depended, however, strongly on assumptions regarding the protection against carriage of the MenACWY vaccine and the CCR: with increasing protection against carriage indirect effects and serotype-replacement increased; alternative assumptions on the CCR (cf. sensitivity analyses D3 and D4) had strong impact on the magnitude of serotype replacement. Based on the alternative CCR from sensitivity analysis D4, e.g., the overall effect of introducing a MenACWY booster at age 16 remained positive, also under an assumption of high protection against carriage, and was larger than the effect reported in the main analysis (SuppFig. 19).

## Discussion

This study aimed to model the long-term effects of introducing an adolescent meningococcal booster program with either MenC or MenACWY conjugate meningococcal vaccines in Germany. Our results showed that both MenC or MenACWY booster vaccinations can prevent additional IMD cases as compared to the status quo (i.e., a no-booster scenario). For the MenACWY booster, our simulations indicated relevant serotype replacement, where the positive effects of prevented ACWY-IMD cases were (partly) offset by an increase in expected IMD cases of other serogroups. This serotype replacement only became noticeable in the simulations 8-10 years after introducing the MenACWY vaccination. The results are generally associated with a high degree of uncertainty around (i) the degree of protection against carriage for the MenACWY vaccine, (ii) future IMD incidence and transmission dynamics, and (iii) the meningococcal carriage prevalence by serogroup in Germany.

The NNV to prevent one IMD case, a case with sequelae or one death were high for all booster scenarios due to low case numbers overall. The uncertainty intervals overlapped for vaccine products in the booster scenarios with a slightly different age of eligibility.

Model results were particularly sensitive to changes in the transmission dynamics, i.e., the future serogroup-specific IMD incidence over the next decade(s), and the degree of protection against meningococcal carriage with MenACWY vaccines. During the COVID-19 pandemic, IMD incidences dropped sharply in Germany and internationally, but the initial post-pandemic data suggest a return to pre-pandemic numbers (19–21). However, various factors, e.g., changes in migration patterns or demographics, may alter (serotype-specific) dynamics in the future. The degree of protection against transmission of the MenACWY vaccine remains unclear, with widely varying results being reported (22, 23).

To date, there is little reliable information on whether MenACWY vaccines lead to serotype replacement in the years following vaccine introduction. Challenges include the low case numbers, the time lag of noticeable replacement effects possibly taking up to 10 years, the potential masking of epidemiological effects by trends in total IMD case numbers, and ongoing changes in vaccination strategies against IMD. Comprehensive, new carriage studies are needed to reliably investigate the current serotype distribution in Germany, and any changes and replacement effects in the future.

For the German context, a modelling study with involvement of a vaccine manufacturer was recently published that reported only a minor impact on IMD incidence and associated mortality after the introduction of a MenC adolescent booster with an assumed uptake of 50%, while the MenACWY adolescent booster was estimated to prevent up to 65 IMD cases per year over a 42-year period (24). The differences to our study can be partly explained by (i) modelling the carriage of meningococcal serogroups independently, which does not rule out co-colonization of individuals by different serotypes, and (ii) larger indirect effects due to a higher assumption of 36% protection against carriage for the ACWY vaccine. In a brief sensitivity analysis assuming no co-colonization, the authors also observed substantial replacement effects.

### Strengths and Limitations

We successfully developed and fit a mathematical model of meningococcal carriage and vaccination to the national surveillance data in Germany in 2005-2019. Previously, such dynamic-transmission models focused on international settings like the UK (10, 12, 25, 26). Our model captured the data and trends in Germany well. For reasons of model parsimoniousness, we fit the model first using serogroup-specific transmission parameters with age-specific scaling of the contact mixing (model 1) before improving the fit with age- and serogroup-specific transmission parameters (model 2). The model can be extended to investigate further (combined) vaccination strategies, and results will inform the discussions of the German NITAG.

We performed various sensitivity and scenario analyses on key parameters and methodological choices. Certain aspects can be regarded as more robust to the changes than others (27), e.g., the average duration of carriage. However, the modelling is more sensitive to changes in the expected age- and serogroup-specific carriage prevalence and case-carrier ratios. The available epidemiological data on the meningococcal carriage prevalence in Germany are sparse and outdated (28). Internationally, the most comprehensive study on the carriage prevalence was from the UK and included data from 1986, 1972 and 1999 (29). In our simulations of the booster vaccination scenarios, we assumed that all individuals with infant immunization received also the adolescent booster vaccine, which can be regarded as an upper-bound estimate of the effectiveness. In reality, the booster uptake could be lower, which we explored with the empirical coverage value of another adolescent vaccination in Germany (against HPV).

### Conclusions

In conclusion, the results of this modelling study show that introducing an adolescent booster vaccination program in Germany is expected to reduce the IMD burden marginally with high NNVs. Although the MenACWY booster vaccines prevent additional IMD cases from the AWY serogroup, the potential protection against carriage may cause substantial serotype replacement in future years that may offset the gain in prevented IMD cases. The meningococcal carriage prevalence and the future IMD incidence remain unclear, and they warrant further monitoring.

## Data Availability

Programming code of the model will be made available upon final publication of the manuscript. Meningococcal IMD case numbers are available from Survstat@RKI (https://survstat.rki.de/).

## Acknowledgements

We thank Dr. Thorsten Rieck for providing estimates of the vaccination uptake against meningococcal disease in Germany.

## Author contributions

Felix Günther: conceptualization; development model structure; model code; formal analysis; data acquisition; initial draft; revision, editing, and approving of the manuscript

Ulrich Reinacher: development model structure; data acquisition; revision, editing, and approving of the manuscript

Sarah Chisholm: development model structure; model code; data acquisition; revision, editing, and approving of the manuscript

Matas Griskaitis: data acquisition; revision, editing, and approving of the manuscript

Michael Höhle: development model structure, supervision; revision, editing, and approving of the manuscript

Stefan Scholz: development model structure; funding; supervision; administration; revision, editing, and approving of the manuscript

Viktoria Schönfeld: formal analysis; revision, editing, and approving of the manuscript

Ole Wichmann: funding; supervision; administration; revision, editing, and approving of the manuscript

Thomas Harder: funding; supervision; administration; revision, editing, and approving of the manuscript

Frank G. Sandmann: conceptualization; development model structure; formal analysis; supervision; administration; initial draft; revision, editing, and approving of the manuscript

