## Supplementary material for "The impact of introducing meningococcal C/ACWY booster vaccination among adolescents in Germany: a dynamic transmission modelling study"

#### Methods and input parameter

##### Supplementary Text S1: Technical details of the dynamic-transmission model

In short, the overall mathematical modelling study followed six steps: First, we built an age-structured deterministic dynamic-transmission model of meningococcal carriage considering multiple serogroups. Second, to connect meningococcal carriage and IMD, we derived age- and serogroup-specific case-carrier ratios from published meningococcal carriage and national surveillance data on IMD cases. Third, we used IMD incidence from national surveillance data by age and serogroup over multiple years to calibrate the dynamic-transmission model to Germany. Fourth, we simulated five vaccination scenarios over 10 years of either continuing the current MenC vaccination in ages 12-23 month (status quo), or additionally introducing an adolescent booster with either monovalent C or polyvalent ACWY vaccines. Fifth, we quantified the effectiveness and efficiency of the adolescent booster in terms of the expected number of IMD cases, sequelae and deaths, and the associated numbers-needed-to-vaccinate (NNVs) to prevent one case of IMD, sequelae or death. Sixth, in sensitivity analyses we explored the impact of assumptions on key parameters and methodological choices.

*ODEs on carriage level*

The dynamic-transmission model of meningococcal carriage describes the transition of individuals between compartments using a set of ordinary differential equations (ODEs). They are defined for the different model compartments as follows:

$$\frac{dS_i(t)}{dt} = -S_i(t) \sum_{X\in\{C,AWY,O\}} \lambda_{i,MenX}(t) + \sum_{X\in\{C,AWY,O\}} \left[r_{MenX}\, Car_{i,MenX}(t)\right],$$

$$\frac{d\, Car_{i,MenX}}{dt} = S_i(t)\lambda_{i,MenX}(t) - r_{MenX}\, Car_{i,MenX}(t),$$

$$\frac{dS_i^{VI,Vac}(t)}{dt} = -S_i^{VI,Vac}(t) \sum_{X\in\{C,AWY,O\}} \left[\left(1-\kappa_{i,MenX}^{Vac}\right)\lambda_{i,MenX}(t)\right] + \sum_{X\in\{C,AWY,O\}} r_{MenX}\, Car_{i,MenX}^{VI,Vac}(t) - \varpi_i^{Vac} S_i^{VI,Vac}(t),$$

$$\frac{dCar_{i,MenX}^{VI,Vac}(t)}{dt} = S_i^{VI,Vac}(t)\left(1-\kappa_{i,MenX}^{Vac}\right)\lambda_{i,MenX}(t) - r_{MenX}\, Car_{i,MenX}^{VI,Vac}(t) - \varpi_i^{Vac} Car_{i,MenX}^{VI,Vac}(t),$$

$$\frac{dS_i^{V,Vac}(t)}{dt} = -S_i^{V,Vac}(t) \sum_{X\in\{C,AWY,O\}} \lambda_{i,MenX}(t) + \sum_{X\in\{C,AWY,O\}} r_{MenX}\, Car_{i,MenX}^{V,Vac}(t) + \varpi_i^{Vac} S_i^{VI,Vac}(t),$$

$$\frac{dCar_{i,MenX}^{V,Vac}(t)}{dt} = S_i^{V,Vac}(t)\ \lambda_{i,MenX}(t) - r_{MenX}\, Car_{i,MenX}^{V,Vac}(t) + \varpi_i^{Vac} Car_{i,MenX}^{VI,Vac}(t),$$

with $i \in \{0, 1, \dots, 84, 85+\}$ being the indicator for 1-year age-groups, $MenX \in \{MenC, MenAWY, MenO\}$ three serogroups considered in the model, and $Vac \in \{MenC, MenACWY\}$ the mono- and polyvalent vaccine. The compartments $S$ and $Car_{MenX}$ represent unvaccinated susceptibles and carriers of the three serogroups, respectively. $S^{VI,Vac}$ and $Car_{MenX}^{VI,Vac}$ represent susceptibles and carriers among individuals vaccinated and currently protected from one of the two vaccines, and $S^{V,Vac}$ and $Car_{MenX}^{V,Vac}$ represent susceptibles and carriers vaccinated individuals, but with waned protection. The combination of three serogroups and two vaccines yields 20 compartments that are defined for each of the 86 age groups (as indicated by subscript $i$). A set of parameters governs the continuous-time transition between compartments:

| | |
|---|---|
| $r_{MenX}$ | = the recovery rate of carriage per serogroup |
| $\varpi_i^{Vac}$ | = the rate of waning of the vaccine-induced protection by age and vaccine type |
| $\kappa_{i,MenX}^{Vac}$ | = the protection against becoming a newly-infected carrier by age, serogroup and vaccine type<br>$\kappa_{i,MenX}^{Vac} \in [0, 1]$ for $Vac \in \{MenC, MenACWY\}$ and $MenX \in \{MenC, MenAWY, MenO\}$, which can vary in the 1-year age-groups $i$. |
| $\lambda_{i,MenX}(t)$ | = the time-varying force of infection by age and serogroup |

Ageing and vaccination are considered via one-year, discrete changes (so-called *step changes*): After integrating the differential equation system over one year, all vaccinations (primary and booster vaccinations) are carried out at a fixed point in time (at the *turn of the year*) and individuals are moved between the compartments correspondingly. Subsequently, all individuals age by one year; technically, the compartments for an age group $i$ are re-initialized by the compartments of age group $i-1$. During the calibration phase on retrospective data, the historical demographic data from Germany is used. At the beginning of a new year, age group $i$, with a new population size according to the known population figures, is initialized by the model-based relative distribution of individuals to the 20 compartments of age group $i-1$ from the end of the previous year and after taking vaccination into account. All newborn individuals in age group 0 are assigned to compartment $S_i(t)$, i.e. treated as susceptible and unvaccinated.

*Force of infection*

The force of infection, $\lambda_{i,MenX}(t)$, is the rate of movement from susceptible compartment, $S_i$, to carrier compartment $Car_{i,MenX}$ of meningococcal serogroup $MenX \in \{C, AWY, Other/B\}$ in age-group $i \in \{0, 1, \dots 84, 85+\}$ at time $t$. It is expressed as a function of currently infected/current carriers of the serogroup $MenX$ in age-groups $j$, contact frequencies between age-groups $c_{i,j}, i, j \in \{0, 1, \dots 84, 85+\}$ specified from the Polymod data (Mossong et al., 2008), and additional parameters estimated from data during the model calibration. The contact frequencies were obtained from the R-package hhh4contacts (Meyer & Held, 2017), contact frequencies are given in 5-year age groups and represent the mean number of daily contacts that an individual from age group $i$ (e.g. 5-9 years old) has with people from age group $j$ (e.g. 50-54 years old). To approximate contact data in a one-year resolution, it was assumed that the contact behavior is identical for all individuals in age group $i$ (e.g. age 5 or 8 years) and that the mean number of daily contacts with persons in age group $j$ (e.g., 50-54 years) is distributed evenly across the one-year age groups (50, 51, ..., 54 years).

Conceptually, the force of infection in our model can be separated into three parts: a serogroup-specific transmission parameter, the rate of contacts between individuals of different age necessary for transmission events to occur, and the proportion of meningococcal carriage in the population for the three groups of serogroups (C, AWY, B/Other). For specifying the age- and serogroup-specific force of infections, we used two different functions/models:

**Model 1a:**

$$\lambda_{i,MenX}(t) = p_{MenX} \sum_{j=0}^{85+} c_{i,j}\zeta_i\zeta_j \frac{Car_{j,MenX}(t) + Car_{j,MenX}^{VI,Vac}(t) + Car_{j,MenX}^{V,Vac}(t)}{N_j(t)}$$

The estimated parameters are:

- $p_{MenX}$, with $MenX \in \{C, AWY, Other/B\}$
- $\zeta_k = \begin{cases} \gamma_1, & 0 \le k \le 4 \\ \gamma_2, & 5 \le k \le 14 \\ \gamma_3, & 15 \le k \le 19 \\ \gamma_4, & 20 \le k \le 85+ \end{cases} \in [0,1]$

The parameters, $p_{MenX}$, correspond to serogroup-specific transmission rates, and $\zeta_k$ are age-specific scaling parameters that down-scale contacts between age-groups, $c_{i,j}$, to contacts relevant for meningococcal transmission. Overall, model 1a has 7 free parameters that are estimated during model calibration.

**Model 1b** is similar to 1a but with 8 age-scaling parameters:

$$\zeta_k = \begin{cases} \gamma_1, & 0 \le k \le 4 \\ \gamma_2, & 5 \le k \le 9 \\ \gamma_3, & 10 \le k \le 14 \\ \gamma_4, & 15 \le k \le 19 \\ \gamma_5, & 20 \le k \le 29 \\ \gamma_6, & 30 \le k \le 49 \\ \gamma_7, & 50 \le k \le 69 \\ \gamma_8, & 70 \le k \le 85+ \end{cases} \in [0,1],$$

corresponding to 11 parameters, overall.

**Model 2:**

In model 2 the age-specific scaling parameters are replaced by age-and serogroup-specific parameter governing transmission rate in age-group i:

$$\lambda_{i,MenX}(t) = p_{MenX,i} \sum_{j=0}^{85} c_{i,j} \frac{Car_{j,MenX}(t) + Car_{j,MenX}^{VI,Vac}(t) + Car_{j,MenX}^{V,Vac}(t)}{N_j(t)}$$

To limit number of free parameters we classify age-groups in 4 (**Model 2a**) or 8 subgroups (**Model 2b**). These are the same as defined in 1a/1b. This corresponds in total to $3 \cdot 4 = 12$ or $3 \cdot 8 = 24$ parameters that are estimated during model calibration.

*Derivation of IMD cases from model*

The dynamic transmission model acts on the carrier level. To derive expected IMD case counts, incident carriers by age, serogroup and year were converted into expected yearly IMD cases using the age- and serogroup-specific case-carrier ratios, CCRs. The CCRs were derived from external data (see below). When deriving the total expected number of IMD cases, the vaccination effect on IMD among the vaccinated and protected new carriers must be considered, incident carriers that are (currently) vaccinated and protect have a reduced probability of developing invasive disease based on the parameter $ve_{i,MenX}^{Vac}$. The overall expected number of IMD cases in age-group $i$, year $k$ and for serogroup $MenX$ is therefore:

$$E\left(IMD_{i,MenX}^{k}\right) = ccr_{i,MenX}\left(NCAR_{i,MenX}^{UV,k} + \sum_{\substack{Vac\,\in[mmCc,\\ pmACWYc]}} NCar_{i,MenX}^{V,Vac,k} + \sum_{\substack{Vac\,\in[mmCc,\\ pmACWYc]}} \left(1 - ve_{i,MenX}^{Vac}\right) NCar_{i,MenX}^{VI,Vac,k}\right),$$

with $NCAR_{i,MenX}^{UV,k}$ the incidence new carriers among unvaccinated, $NCar_{i,MenX}^{V,Vac,k}$ among vaccinated with waned protection, and $NCar_{i,MenX}^{VI,Vac,k}$ among vaccinated and currently protected individuals in age-group $i$, year $k$, vaccine type $Vac$ and serogroup $MenX$.

*Case-carrier ratio*

The CCRs were estimated based on external data the following way: First, we used published point-prevalence data from individuals aged 3-26 years in Germany in 1999-2000 (Claus et al., 2005), which we supplemented with data from adults in Europe in 2010 and from individuals aged 65+ years in Germany in 2019 (Christensen et al., 2010; Drayss et al., 2019). The latter two studies where operationalized as a sero-prevalence of 13% in 30-year old, 8% in 50-year old and 0.4% in 70-year old, cumulative over all serogroups. From the (Claus et al., 2005) study, we extracted case numbers of MenC, MenAWY, and Men Other/B carriers among tested individuals in 1-year age groups and estimated an age- and serogroup-specific carriage prevalence from these data-points based on a generalized quasi-Poisson linear model with non-linear age-effect and fixed serogroup effect and the log-number of tested individuals as offset. Carriage prevalence was estimated highest in adolescents and young adults, with a peak around age 20 years. The group of meningococcal serogroups “Other/B” showed the highest magnitude in the prevalence, followed by the serogroups AWY and then C (Supplementary Figure ST1 A).

Assuming a mean duration of carriage of 6 month, we derived an expected yearly carrier incidence by multiplying the estimated prevalence with factor 2.

As a second data source, we also obtained the average yearly age-and serogroup-specific IMD incidence of the years 2002 to 2005 (Supplementary Figure ST1 B).

To estimate the case-carrier ratio for individuals aged 3 to 70 years, we again used a quasi-Poisson GAM with the IMD incidence as the outcome, a smooth age and a fixed serogroup effect as covariates and the log-number of new carriers per year as an offset, derived by multiplying the estimated annual carrier incidence by the mean population size by age in 2002-2005.

For our main analysis, we kept the CCRs constant outside this range (i.e. for the age groups 0-2 and 70-85+).

The combination of a low carriage prevalence and comparatively high IMD incidence resulted in estimated CCRs that were highest for infants and young children before drastically decreasing into adolescent age and further decreasing for adults before increasing again in older age (Supplementary Figure ST1 C). Similarly, the combination of a high prevalence of carriers but a comparatively low incidence of IMD lead to the highest CCR in serogroup C, followed by B/Other and the lowest CCR for serogroup AWY.

Since the CCR also has a direct influence on the initialization of the compartments and thus on the assumed carriage prevalence during calibration and simulation, we performed two sensitivity analyses regarding the CCR. In a first sensitivity analysis, we used the same data sources a s in the main analysis but two alternative analysis choices: First, instead of restricting the analysis to the age range of 3-70 years (due to a lack of data on carriage in Germany outside of this age range) and extrapolating the CCR for younger/older persons outside of this age range as constant, we have instead assumed the carriage prevalence in these age ranges constant and determined the CCR from the derived yearly carriage incidence and the actual (mean) IMD case numbers for age 0-2 and 71-85+. Second, we estimated the CCR from the IMD case numbers and carriage incidence based on serogroup-specific non-linear age-effect instead of using an overall non-linear age-effect and serogroup-specific fixed effect. For the second sensitivity analysis we used the published data on estimated age-specific carriage from the systematic review of (Christensen et al., 2010), and distributed the to the three serogroups considered based on the relative share of serogroups from (Claus et al., 2005) aggregated over all ages (3-26 years). Compared to the main analysis, the main implications of the approach in the first sensitivity analysis were a substantially increased CCR in all serogroups for the age range 0-2 years, since the higher IMD figures in this age range are now not reflected by higher carriage at constant CCR, but rather by a higher CCR at constant carriage. Furthermore, a slightly higher CCR was derived for the 70 to 80-year age group in the AWY serogroup (Supplementary Figure ST2 B). Based on the data of (Christensen et al., 2010), a higher carriage prevalence was assumed in age groups 0–15 years and 50+ in sensitivity analysis 2 compared to the main analysis, which in turn led to significantly lower CCRs, especially at a young age (Supplementary Figure ST2 A and B).

*Supplementary Figure ST1: Estimated meningococcal carriage prevalence by age and serogroups in Germany (panel A), the observed IMD incidence by age and serogroups in Germany in 2002-2005 from national surveillance, SurvStat (panel B); the estimated case-carrier ratio by age and serogroups based on the carriage prevalence and the IMD incidence in Germany in 2002-2005 (panel C). Dots represent observed data (panel A: point prevalence, panel B: mean incidence, panel C: ratio of estimated yearly carrier incidence and observed IMD incidence); solid lines show model-based smooth estimates.*

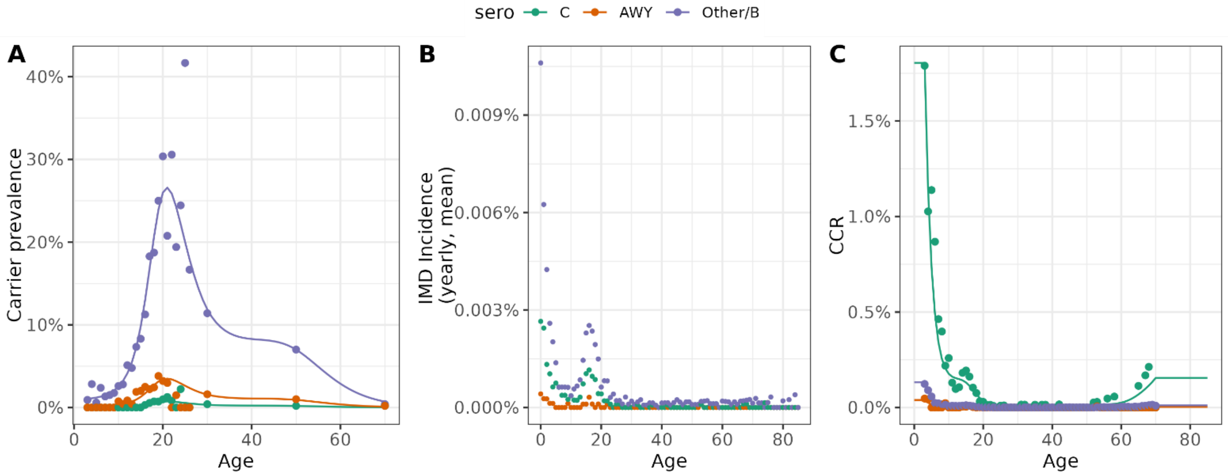

*Supplementary Figure ST2: Sensitivity analyses CCR. Carrier prevalence (panel A) and derived CCR panel B) by serogroup for the main analysis and two sensitivity analyses based on different methodological choices and data sources.*

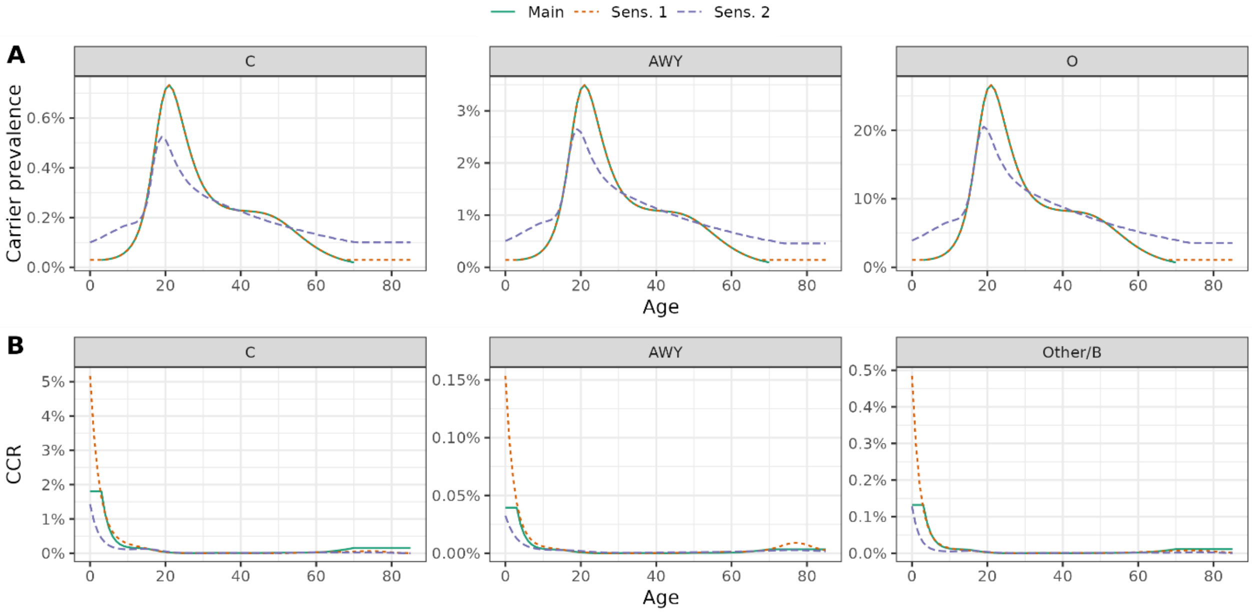

*Derivation of sequelae and fatal cases*

We used published estimates of the probabilities of 16 different long-term health outcomes (Scholz et al., 2019) to derive estimated cases with (at least one) sequelae from the IMD case numbers, assuming that the different outcomes occur independently of each other. We also assumed that they occurred independently of the age of the IMD cases. The resulting probability of at least on sequelae was 34.8%.

For deaths we estimated the case-fatality ratio (CFR) in Germany by age and serogroup using national surveillance data from 2002-2019 with complete records for age, sex, serogroup and cause of death (confirmed IMD, n=6654). We used a binomial generalized additive model to estimate the number of deaths among all IMD cases with age and serogroup (C, AWY, B/Other) as independent variables. Using separate smoothing splines for age by serogroup and an additive effect of serogroup, the estimated CFRs were specific to age and serogroup in Germany. The estimated CFRs for serogroups C and AWY were concave and highest for adults aged 40-50 years before declining to higher levels in older individuals than for children and young adults, while for serogroup B/Other the CFRs were convex with a minimum around age 20 years and again higher in old age than for children and adolescents (Supplementary Figure ST3).

We quantified the effect of different vaccination strategies by reporting the expected number of prevented IMD cases, the expected number of prevented IMD cases with sequelae or death, and the expected number of prevented deaths due to IMD compared to the baseline vaccination strategy (status quo). To compare the efficiency of different strategies, we also derived corresponding NNVs. Per age- and serogroup, we derived the prevented IMD cases with sequelae or death as $\#IMD \cdot (CFR + (1 - CFR) \cdot CSR)$ and prevented deaths as $\#IMD \cdot CFR$, where $\#IMD$ is the expected number of prevented IMD cases, $CFR$ is the age- and serogroup-specific case fatality ratio, and $CSR$ is the probability of developing at least one sequelae among surviving IMD cases (assumed constant by age and serogroup).

*Supplementary Figure ST3: Estimated meningococcal case fatality ratio based on national surveillance data from 2002-2019.*

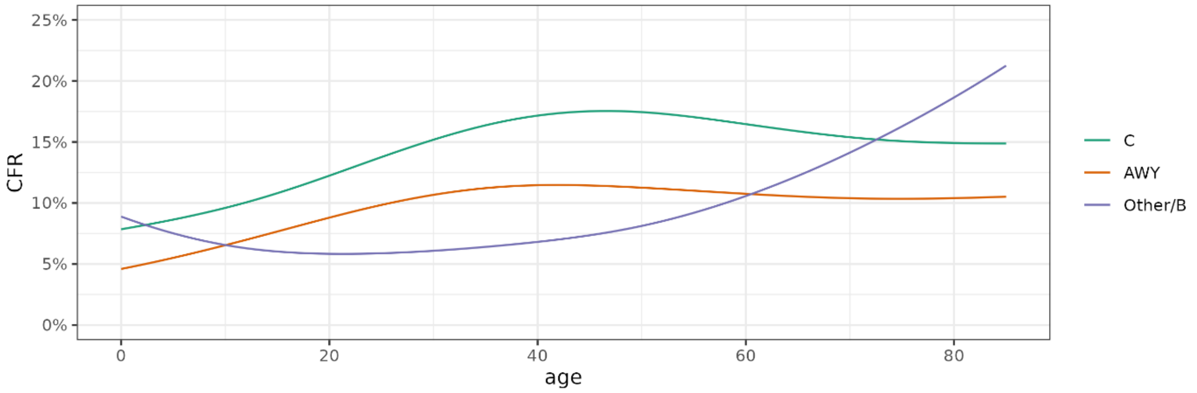


*Initialization of compartments for the calibration and simulation period*

We initialized the compartments (susceptibles and carrier of MenC/AWY/O) of the dynamic transmission model at the start of the calibration period (01.01.2005) based on the sero- and age-specific mean yearly IMD cases from the years 2002-2005. These mean incident IMD cases where transformed to age- and serogroup-specific incident carriers by dividing through the estimated CCRs. To obtain a smooth estimate of (relative) age- and serogroup-specific carriage prevalence, we multiplied the derived incident (yearly) carriers with the mean duration of carriage (0.5 years in the main analysis) and used these synthetic case counts as outcome of a Poisson generalized additive model with log population-size as offset and smooth effects of age for each serogroup. The resulting age- and serogroup-specific carriage prevalence estimates where used for initialization of the model. All individuals where placed in the unvaccinated compartments as the calibration period started before the start of the MenC infant immunization program in Germany.

Our simulation period directly followed the calibration period, and we initialized the DTM to simulate the vaccination effects based on the relative distribution of the population to the different compartments (susceptible and carriage of the 3 serogroups, among unvaccinated, vaccinated and protected, as well as vaccinated without vaccine protection) at the end of the calibration period.

*Model fitting*

For each of the four models (1a-b, 2a-b), we estimated parameters via Maximum Likelihood as follows: Each model was initialized as described. Afterwards, the model was numerically solved for a given set of parameters for 2005-2019. The time was chosen to cover the period of the MenC primary vaccination introduction in 2006 (STIKO, 2006), and up until the start of the COVID-19 pandemic in 2020. The model also considered the increasing MenC vaccination coverage during the calibration period. The derived number of incident carriers by age, serogroup and year were transformed into expected yearly IMD cases using the estimated CCRs. Then, assuming a Negative Binomial distribution of the yearly IMD cases by age and serogroup observed in the surveillance data, we optimized the model for the unknown parameters by numerical minimization of the negative log-likelihood using a parallel implementation of the L-BFGS-B algorithm (Gerber & Furrer, 2019). In addition to the free parameters governing the force of infection per model, we estimated an additional parameter capturing the overdispersion of the Negative Binomial distribution compared to the Poisson distribution. Following standard maximum likelihood theory, parameter uncertainty was quantified based on the inverse Fisher information matrix (Hessian) evaluated at the minimum of the negative log-likelihood that corresponds to the variance-covariance matrix of the asymptotically Multivariate normal distributed Maximum Likelihood estimator. Uncertainty was propagated through the modelling and simulation by sampling 100 parameter vectors from this Multivariate Normal distribution, solving the ODE model, as well as calculating quantities of interest (e.g., expected case counts or differences in expected case counts) for each parameter combination, and calculation of uncertainty intervals based on empirical quantiles of the resulting distributions.

Model selection among the four models was performed against Akaike and Bayesian information criteria (AIC, BIC). The results from the best-fitting model were used for simulations in the main analysis, and from the second-best-fitting model in sensitivity analyses.

**Supplementary Text S2: Model input data and operationalization**

*Input data*

To parameterize the model, we used published data from the literature, performed analyses of external data and performed a model calibration (cf. Supplementary table S1 for an overview of the data sources). To calibrate the model, we used national surveillance data of notifiable IMD cases by age and serogroup in Germany in 2005-2019. Cases with unknown serotype (between 8.8% and 16.0% per year) were distributed to the three serogroups considered (C, AWY, Other/B) according to the relative serogroup distribution in the respective year. Routinely collected data on vaccine coverage were used for the primary vaccination during the fitting and to inform the future vaccination scenarios, too (‘KV-Impfsurveillance’) (Robert Koch-Institut, 2024). For the case-carrier ratio (CCR) we used published point-prevalence data on meningococcal carriage from individuals aged 3-26 years in Germany in 1999-2000 (Claus et al., 2005), which we supplemented with data from adults in Europe in

2010 and from individuals aged 65+ years in Germany in 2019 (Christensen et al., 2010; Drayss et al., 2019) (cf. Supplementary Text S1: Case-carrier ratio).

For the mean duration of carriage, we assumed 6 months (Christensen et al., 2016) and 12 months in a sensitivity analysis (De Wals & Bouckaert, 1985). For the contact rates in the force of infection, we used empirical contact data in Germany from the POLYMOD study (Mossong et al., 2008). Demographic data for Germany in 2005-2019 were used during the calibration (Statistisches Bundesamt Destatis 2023a), for the forwards simulations we used the available data for 2020-2022 and the official population projections for 2023-2049 (Statistisches Bundesamt Destatis 2023b).

Information on vaccine effectiveness and the duration of the vaccine protection were collected in a systematic review (Griskaitis et al., 2024). For modelling, we mainly utilized published estimates on the vaccine effectiveness against IMD from real-world post-licensure surveillance data in the UK (Campbell et al., 2010). A detailed description and discussion of our assumption is given in the following subsection.

For the derivation of expected case numbers with sequelae and fatal cases from expected IMD cases, we used published estimates of the probabilities of 16 different long-term health outcomes (Scholz et al., 2019) and analyzed German surveillance data from 2002-2019 (cf. Supplementary Text S1: Derivation of sequelae and fatal cases).

*Protection from vaccination*

Data on vaccine efficacy and effectiveness of the mono- and polyvalent MenC/MenACWY vaccines as primary immunization in infants or adolescent booster vaccine are scarce, in particular with respect to protection against carriage and duration of protection. Due to the relatively low IMD incidence, many studies on vaccine efficacy or effectiveness, including licensure trials, were based on immunogenicity data (correlates of protection). There exist few controlled trials with clinical endpoints and few estimates of vaccine effectiveness from observational studies (Griskaitis et al., 2024).

For the monovalent MenC vaccine, real-world VE against IMD was tracked during post licensure surveillance after introducing a routine infant immunization program in 1999. (Campbell et al., 2010) reported real-world VE estimates against IMD for the routine infant immunization program (3-dose schedule during first year of life) in UK based on the screening method: during the first year since vaccination, VE against MenC-IMD was estimated as 96% (95%-CI: 87-99%); VE point estimates decreased to 68%, 60%, and 31% for year 2, 3, and 4-5 after vaccination, respectively. Estimated MenC-VE was similar for a one-dose catch-up in toddlers between 12-23 month of age (VE: 89%; 95%-CI: 64-98% during first year after vaccination) and high in children and adolescents (VE in 3-18 year old during first year after vaccination: 96%; 95%-CI: 92-99%; slightly less reduction in VE for longer observation periods compared to infants/toddlers). (Andrews et al., 2003) reported an observed VE against MenC-IMD in toddlers between 12-23 month of age of 90.1% (95%-CI: 74.9-96.1%) during an observation period with average time since vaccination of 10 month.

With respect to protection against C-carriage, a VE estimate of 75% (95%-CI: 23-92%) in the UK was reported by (Maiden et al., 2008) based on data from a multi-center meningococcal carriage study in 15-19-year-old students with self-reported vaccination status of MenC carriers from the 2001 wave of the study (2 years after introduction of the immunization program).

For the polyvalent MenACWY vaccine real-world estimates of VE against IMD using the screening method were estimated during times of MenW outbreaks in Great Britain, Australia and the Netherlands for different Follow-up periods (up to 4 years, 2-3 years, ~2.5 years) and different target groups (adolescents in GB and infants/young children in Australia and Netherlands) (Campbell et al., 2022; Ewe et al., 2024; Ohm et al., 2022). All estimates show high protection with VEs against ACWY-

IMD in the range between 80% and 94% depending on the serotype, study and target groups and are associated with substantial uncertainty.

Potential protection against carriage and its degree is controversial for the MenACWY vaccine. On their vaccine information website, for example, the UKHSA state "This [MenACWY] vaccine will provide direct protection to the vaccinated cohort and, by reducing carriage of MenW, will also provide indirect protection to unvaccinated children and adults" (UKHSA, 2015), while the CDC writes "[US surveillance] data suggest MenACWY vaccines have provided protection to those vaccinated, but not to the larger, unvaccinated community through population or herd immunity" (CDC, 2023). Based on data from a randomized, observer-blind, controlled phase 3 trial in University students in England, (Read et al., 2014) reported (cumulative) new acquisition of CWY carriage among 59 of 712 MenACWY vaccinated persons and 68 of 709 placebo vaccinated persons at time-points 1, 3, 5, or 11 months after second vaccination. This corresponds to a RR of 0.86 (95%-CI: 0.61-1.22) and a corresponding VE estimate against MenACWY carriage of 0.14 (95%-CI: -0.22 – 0.39) during the first year after MenACWY vaccination (own calculations). (Carr et al., 2022) reported substantially lower MenCWY carriage in a cross-sectional study of students postimplementation of the MenACWY vaccination program in UK school students (2018, n=13,438; 15-19 years) compared to a similar independent cross-sectional carriage study pre-implementation (2014-2015, n=10,625) of the vaccination program (MenCWY carriage prevalence 0.71% compared to 2.03% pre-implementation, OR=0.34; 95%-CI: 0.27-0.44). This can be interpreted as evidence for indirect effects of the vaccination program, but the reported OR is difficult to be interpreted as a measure of vaccine effectiveness, since differences in observed carriage prevalence might be influenced by direct effects of vaccination, as well as indirect effects, and/or general time or cohort effects. No individual-level information on vaccination status of the study participants was reported.

In our model, vaccination is operationalized through vaccine-specific compartments for individuals protected from the respective vaccine (cf. Supplementary Text S1). Vaccinated individuals that are currently part of this compartment have a reduced rate of becoming a MenC or MenAWY carrier compared to unvaccinated individuals (specified based on a parameter reducing the force of infection among vaccinated and protected) and move out of the vaccine protected compartment into a vaccine waned compartment based on a fixed (age-specific) rate (exponential waning of vaccine induced protection). Furthermore, the CCR for IMD among new carriers is reduced by a multiplicative factor. In the following, we describe specification of the three parameters of the VE model based on the available data described above.

As data on the real-world vaccine effectiveness for the mono- and polyvalent vaccines are rare, and do not point towards substantial differences in protection, we assume the same overall VE against IMD for the MenC vaccine and the MenACWY vaccine for our main analysis. Note, however that the monovalent vaccine only protects against MenC-IMD and the polyvalent against MenC- and MenAWY-IMD. To specify parameters of the VE model in the main analysis, we fitted an exponential waning model with two parameters (initial VE and mean duration of protection) to the time-specific data on VE against MenC-IMD in infants from (Campbell et al., 2010) using least squares. The resulting estimate pointed towards an initial VE of 1 against IMD and a mean duration of protection of 4 years (exponential waning with rate=1/4). The additional parameter for protection against MenC carriage as well as and MenAWY carriage for the polyvalent vaccine were specified based on an assumed 1-year carriage-VE of 75% for the MenC vaccine and a 1-year carriage-VE of 14% for MenACWY based on the data from (Maiden et al., 2008) and (Read et al., 2014), respectively. Since data from real-world VE and immunogenicity data point towards longer-lasting immune response in adolescents compared to infants, we increase the mean duration of protections for individuals >=12 years from 4 to 10 years, this age-group corresponds also to the eligible age-group for booster vaccination in our simulation scenarios. All VE-related parameters for our model in the main analysis are shown in Supplementary Table 1.

**Supplementary Table 1: model parameter and data sources**

| Type | Description/Value | Reference(s) |
|---|---|---|
| IMD case numbers used for calibration | IMD case numbers by age and serogroup from years 2005-2019, derived from national surveillance data (Survstat@RKI) | Survstat@RKI |
| Vaccination numbers used for calibration | Vaccination numbers collected at RKI (KV-Impfsurveillance) | (Robert Koch-Institut, 2024) |
| Case-carrier ratio (CCR) | Age- and serogroup-specific CCR derived from cross-sectional carriage data and IMD incidence in Germany. | Carriage data:<br>(Christensen et al., 2010; Claus et al., 2005; Drayss et al., 2019)<br>IMD incidence 2002-2005: Survstat@RKI<br>Details on derivation and operationalization in SuppText. *S1: Case-carrier ratio* |
| Mean duration of carriage | 6 months (Sens. Analysis: 12 month) | (Christensen et al., 2016; De Wals & Bouckaert, 1985) |
| Force of infection (FOI) | Age- and serogroup-specific FOI estimated during model calibration. | Details in *SuppText. S1: Force of infection* |
| Vaccine efficacy | Parameters from fitting an exponential waning model to data on real-world vaccine efficacy:<br>- Protection against carriage (among vaccinated and protected):<br>C: 0.85/0.0/0.0 vs. MenC/AWY/Other<br>ACWY: 0.16/0.16/0.0 vs. MenC/AWY/Other<br>- Protection against IMD among (new) carriers:<br>C: 1.0/0.0/0.0 vs. MenC/AWY/Other<br>ACWY: 1.0/1.0/0.0 vs. MenC/AWY/Other<br>- Duration of vaccine-induced protection:<br>C and ACWY: 4 years up to age 12, 10 years afterwards<br>- Corresponding total protection against C or ACWY IMD:<br>88%, 69%, 54%, 38% (during year 1, 2, 3, and 4-5 after vaccination)<br>- Corresponding protection against C/ACWY carriage in first year after vaccination:<br>MenC: 75.2%, MenACWY: 14.2% | Real-world VE estimates from post licensure surveillance after introducing routine infant immunization program in UK: (Campbell et al., 2010)<br><br>Details and further references in *Supp. Text S2*. |
| Demographic data | Official population data by Destatis up until 2022 and population projections for simulations up until 2049 | (Statistisches Bundesamt Destatis 2023a, 2023b) |
| Contact frequencies | Polymod | (Mossong et al., 2008) |
| Sequelae and fatalities among IMD cases | Published estimates of the probabilities of 16 different long-term health outcomes for sequelae and national surveillance data from 2002-2019 (Survstat@RKI) for fatalities. | (Scholz et al., 2019)<br>Survstat@RKI<br>Details in *SuppText. S1* |

### Results

#### Model calibration

*Supplementary Table 2A: model selection*

| *Model* | *AIC* | *BIC* |
|---|---|---|
| *2b* | 9,404 | 9,561 |
| *1b* | 9,576 | 9,651 |
| *2a* | 10,556 | 10,638 |
| *1a* | 10,570 | 10,621 |

*Supplementary Table 2B: Fitted parameters model 2b. Est. is the point estimate, SE the associated standard error from fitting the model during calibration. Parameters p are serogroup and age-specific transmission rates, φ is the overdispersion parameter of the negative binomial distribution with mean* $\mu$ *and variance* $\mu + \varphi * \mu^2$.

| | est | se |
|---|---|---|
| $p_{C,\ 0-4}$ | 0.027 | 0.0026 |
| $p_{C,\ 5-9}$ | 0.027 | 0.0044 |
| $p_{C,\ 10-14}$ | 0.086 | 0.0103 |
| $p_{C,\ 15-19}$ | 0.231 | 0.0145 |
| $p_{C,\ 20-29}$ | 0.566 | 0.0265 |
| $p_{C,\ 30-49}$ | 0.307 | 0.0163 |
| $p_{C,\ 50-69}$ | 0.140 | 0.0122 |
| $p_{C,\ 70-85+}$ | 0.050 | 0.0067 |
| $p_{AWY,\ 0-4}$ | 0.011 | 0.0016 |
| $p_{AWY,\ 5-9}$ | 0.010 | 0.0031 |
| $p_{AWY,\ 10-14}$ | 0.048 | 0.0101 |
| $p_{AWY,\ 15-19}$ | 0.218 | 0.0196 |
| $p_{AWY,\ 20-29}$ | 0.632 | 0.0402 |
| $p_{AWY,\ 30-49}$ | 0.269 | 0.0285 |
| $p_{AWY,\ 50-69}$ | 0.183 | 0.0182 |
| $p_{AWY,\ 70-85+}$ | 0.144 | 0.0160 |
| $p_{Other,\ 0-4}$ | 0.039 | 0.0027 |
| $p_{Other,\ 5-9}$ | 0.038 | 0.0034 |
| $p_{Other,\ 10-14}$ | 0.058 | 0.0054 |
| $p_{Other,\ 15-19}$ | 0.209 | 0.0115 |
| $p_{Other,\ 20-29}$ | 0.600 | 0.0255 |
| $p_{Other,\ 30-49}$ | 0.282 | 0.0125 |
| $p_{Other,\ 50-69}$ | 0.136 | 0.0081 |
| $p_{Other,\ 70-85+}$ | 0.055 | 0.0045 |
| $\varphi$ | 0.253 | 0.0234 |

*Supplementary Figure 1: detailed model fit: black dots are the model based, expected yearly IMD cases per sero- and age-group (columns and rows), the error bars are the corresponding 95%-UIs. Ribbons represent the 2.5%- and 97.5%-quantiles of the predictive Poisson distribution. Bars correspond to reported case counts from surveillance data. Shown are the results for the best fitting model (2b).*

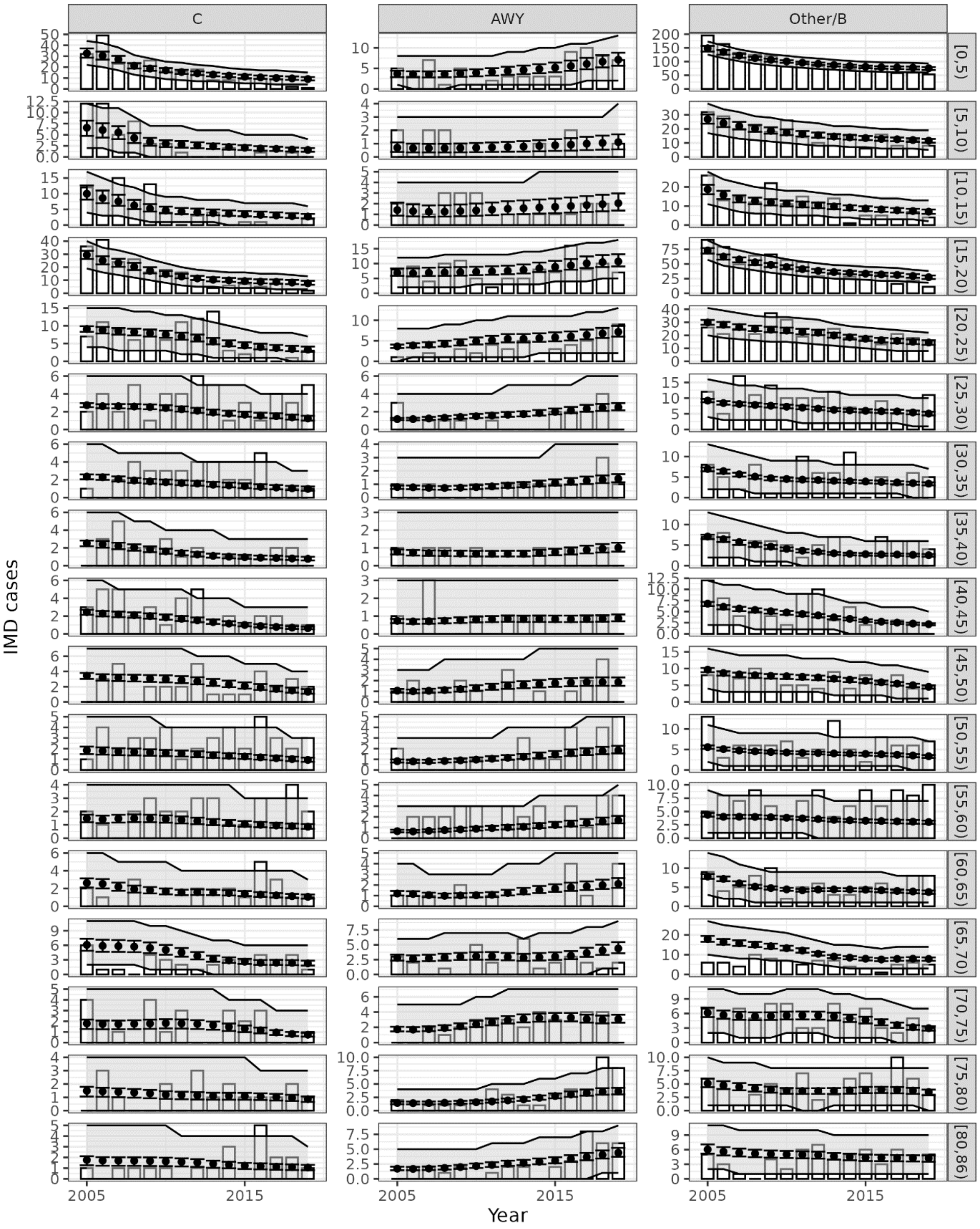

*Supplementary Figure 2: age-specific carriage prevalence by serogroup during calibration period in fitted model*

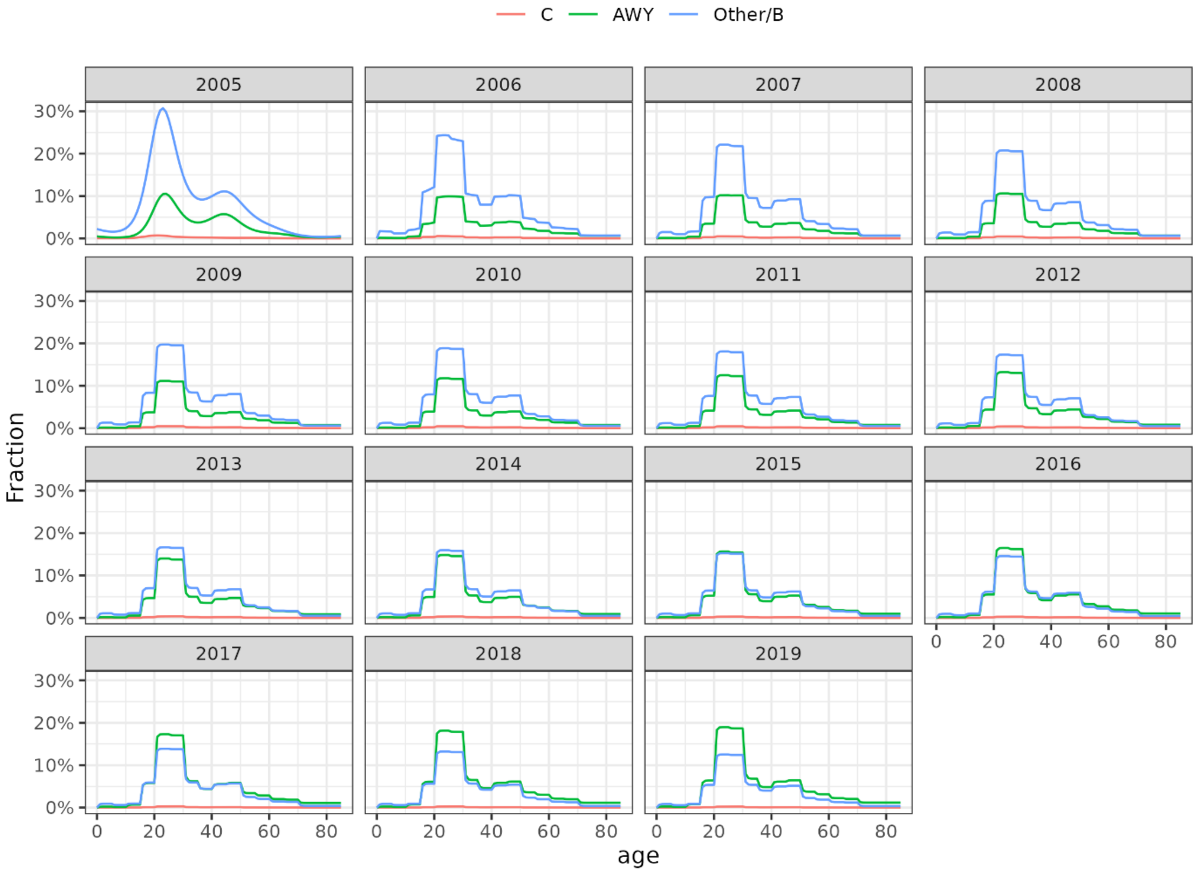

#### Simulation of adolescent booster strategies

*Supplementary Figure 3: age-specific carriage by serogroup (rows) in selected years of simulation period (columns) for simulation scenarios 1-3 (no booster, MenC, MenACWY at age 13)*

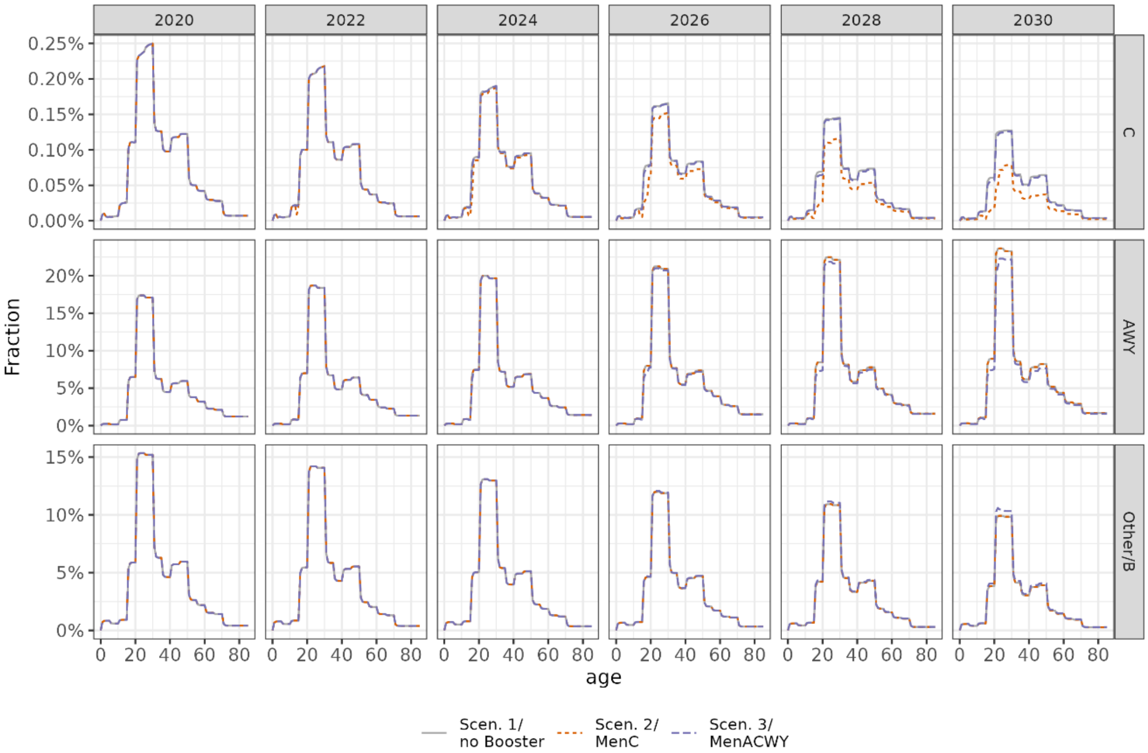


#### Modelled total effectiveness and efficiency of introducing the adolescent booster

*Supplementary Table 3: Serogroup-specific changes in expected outcomes (prevented cases) for the different adolescent booster scenarios compared to simulation scenario 1 (no booster)*

| | | IMD | | | Sequelae or Death | | | Death | | |
|---|---|---|---|---|---|---|---|---|---|---|
| Scenario | Serogroup | Prevented | 2.5%-UI | 97.5%-UI | Prevented | 2.5%-UI | 97.5%-UI | Prevented | 2.5%-UI | 97.5%-UI |
| | C | 51,2 | 40,0 | 67,8 | 21,7 | 17,0 | 28,8 | 6,0 | 4,7 | 7,9 |
| 2 | AWY | -0,3 | -0,4 | -0,2 | -0,1 | -0,2 | -0,1 | 0,0 | 0,0 | 0,0 |
| | Other/B | -0,5 | -0,7 | -0,3 | -0,2 | -0,3 | -0,1 | 0,0 | -0,1 | 0,0 |
| | C | 24,6 | 19,4 | 32,4 | 10,3 | 8,1 | 13,6 | 2,7 | 2,1 | 3,6 |
| 3 | AWY | 66,0 | 56,6 | 85,1 | 26,5 | 22,7 | 34,1 | 5,3 | 4,6 | 6,9 |
| | Other/B | -13,8 | -18,0 | -10,3 | -5,5 | -7,2 | -4,1 | -1,1 | -1,4 | -0,8 |
| | C | 81,4 | 62,3 | 106,7 | 34,7 | 26,5 | 45,5 | 9,7 | 7,4 | 12,8 |
| 4 | AWY | -0,7 | -0,9 | -0,5 | -0,3 | -0,4 | -0,2 | -0,1 | -0,1 | 0,0 |
| | Other/B | -1,1 | -1,5 | -0,7 | -0,5 | -0,6 | -0,3 | -0,1 | -0,1 | -0,1 |
| | C | 31,0 | 22,8 | 40,7 | 13,1 | 9,6 | 17,2 | 3,6 | 2,6 | 4,7 |
| 5 | AWY | 92,6 | 81,1 | 114,8 | 37,4 | 32,8 | 46,3 | 7,9 | 6,9 | 9,7 |
| | Other/B | -40,3 | -49,0 | -32,7 | -16,1 | -19,6 | -13,1 | -3,3 | -3,9 | -2,6 |

*Supplementary Table 4: Model results of the effectiveness and efficiency of introducing the adolescent booster vaccination program in Germany over 10 years excluding other/B serogroup (as compared to no-booster program, scenario 1).*

| | | **Prevented total number of outcomes (95%-UI)** | | | **Numbers needed to vaccinate to prevent one outcome (95%-UI)** | | |
|---|---|---|---|---|---|---|---|
| **Scenario** | **Outcome** | **Mean** | **Lower 95%-UI** | **Upper 95%-UI** | **Mean** | **Lower 95%-UI** | **Upper 95%-UI** |
| 2 | IMD cases | 51 | 40 | 67 | 140,000 | 100,000 | 170,000 |
| | Sequalae or death | 22 | 17 | 29 | 320,000 | 240,000 | 410,000 |
| | Death | 6 | 5 | 8 | 1,200,000 | 880,000 | 1,500,000 |
| 3 | IMD cases | 91 | 80 | 110 | 77,000 | 64,000 | 87,000 |
| | Sequalae or death | 37 | 32 | 44 | 190,000 | 160,000 | 210,000 |
| | Death | 8 | 7 | 10 | 860,000 | 730,000 | 980,000 |
| 4 | IMD cases | 81 | 62 | 110 | 83,000 | 63,000 | 110,000 |
| | Sequalae or death | 34 | 26 | 45 | 190,000 | 150,000 | 250,000 |
| | Death | 10 | 7 | 13 | 690,000 | 530,000 | 910,000 |
| 5 | IMD cases | 120 | 110 | 140 | 54,000 | 46,000 | 61,000 |
| | Sequalae or death | 50 | 45 | 59 | 130,000 | 110,000 | 150,000 |
| | Death | 11 | 10 | 13 | 580,000 | 510,000 | 660,000 |

#### Sensitivity analyses of simulation results

##### Supplementary Text S3: Results sensitivity analyses

We explored the impact of key parameters and methodological choices on the simulation of the effectiveness and efficiency of the booster vaccination. In the following, we summarise results based on the different types of parameters/assumptions varied. Supplementary Figure 4 shows the overall results of all sensitivity analyses for the booster vaccines in life year 16 in the same way as for the booster in life year 13 in Figure 5 of the main text. Supplementary Figures 5-15 summarize extrapolated case counts and overall effects of the booster vaccines for the sensitivity analyses in the same way as in Figure 3 of the main text for the main analysis.

*Supplementary Figure 4: Results* of the sensitivity analyses for booster vaccination in life year 16: Expected IMD cases during the simulation period without booster vaccination for each (sensitivity) analysis and relative change compared to main analysis (panel A); prevented expected IMD cases based on the booster vaccination program in life year 16 (C or ACWY vaccine, color coded) for each (sensitivity) analysis and relative change compared to prevented cases in the main analysis (panel B); corresponding NNVs and relative change compared to main analysis (panel C).

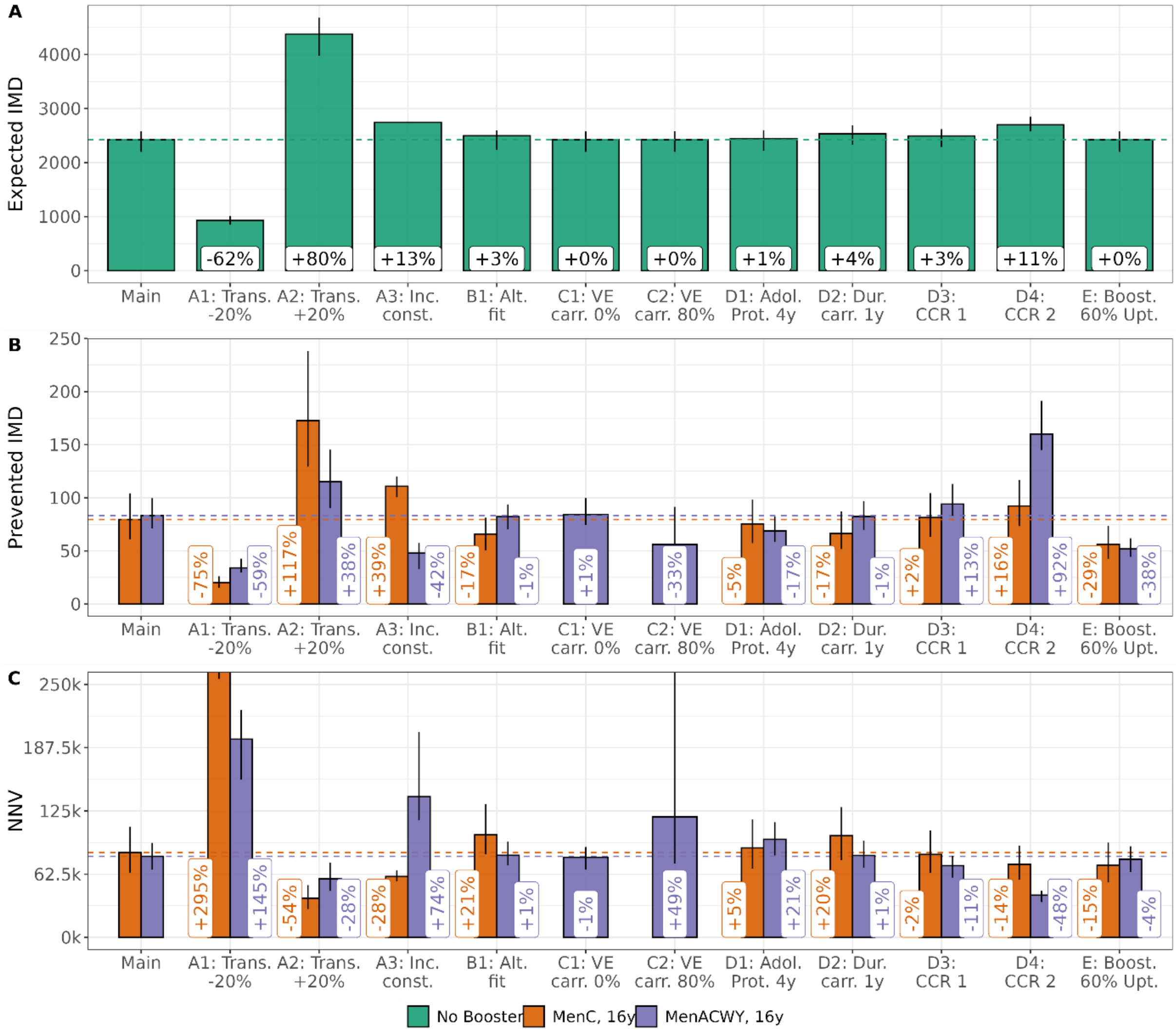

*S3 A: Transmission dynamics during extrapolation.*

The sensitivity analyses revealed that results were sensitive to the transmission dynamics assumed during the simulation period, with smaller (serotype-specific) transmission and expected IMD case counts reducing effectiveness and efficiency of the booster vaccinations (A1), while higher expected case counts led to higher effectiveness and efficiency (A2; Figure 5 for booster in life year 13, SuppFig. 4 for booster in life year 16, SuppFig. 5-6). Keeping the incidence constant (A3) increased effectiveness and efficiency of the MenC booster scenario, and reduced efficiency of the MenACWY scenario as expected case numbers of MenC-IMD was considerably increased and expected case numbers of MenAWY-IMD decreased compared to the main analysis (SuppFig. 7).

*Supplementary Figure 5:* ***Expected number of IMD cases by serogroup in each vaccination scenario 1-5 over 10 years, 2020-2029, sensitivity A1 (reduced transmission)****. Cumulative total number of cases and relative change compared to scenario 1 (panel A), annual number of cases by serogroup (panel B) and relative change in the cumulative number of cases by sero- and age-group (panel C).*

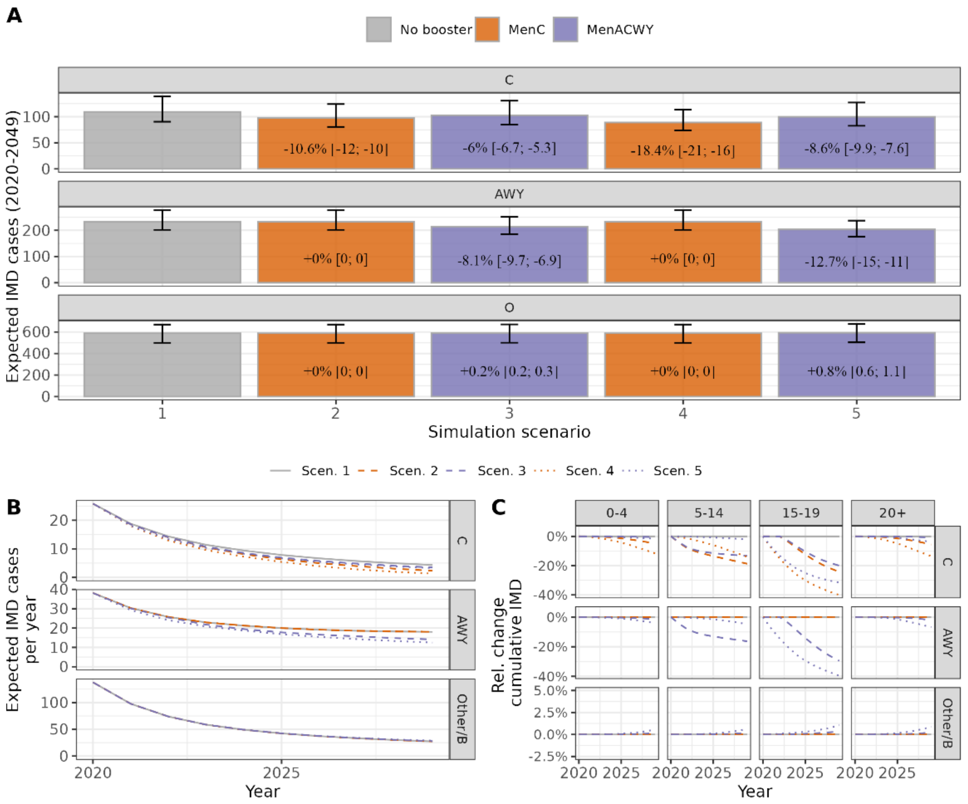


Expected case numbers and absolute numbers of prevented cases are considerably decreased compared to main analysis, leading to a lower overall effectiveness and efficiency of the booster vaccination as compared to the main analysis.

*Supplementary Figure 6:* ***Expected number of IMD cases by serogroup in each vaccination scenario 1-5 over 10 years, 2020-2029, sensitivity A2 (increased transmission).***

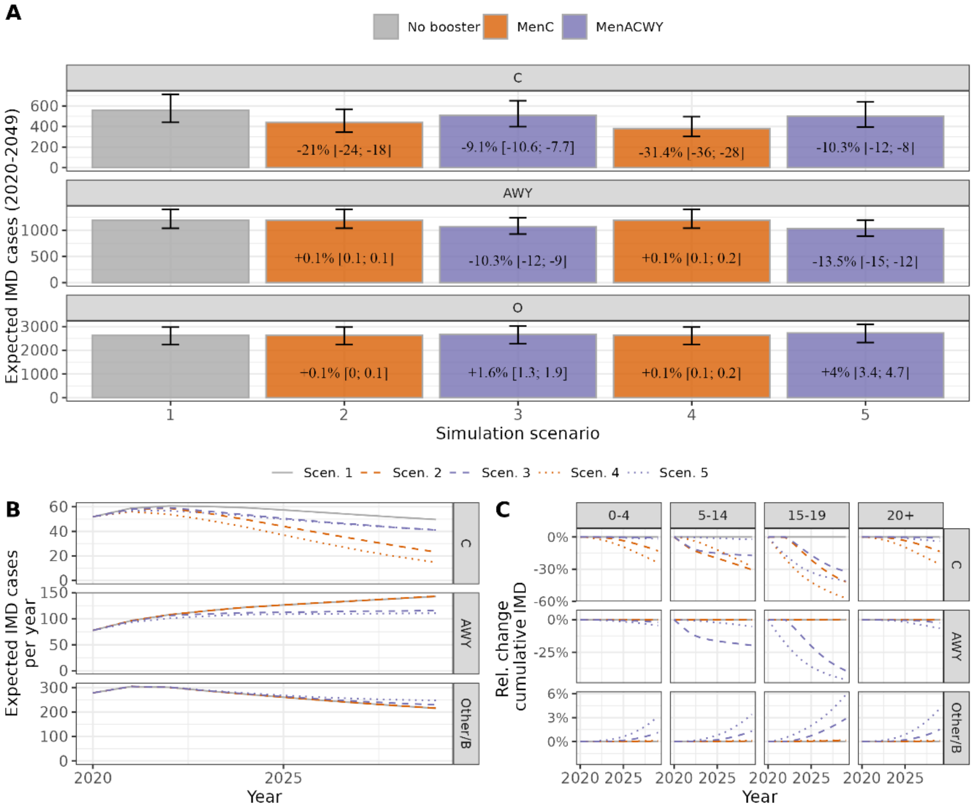


Expected case numbers and absolute numbers of prevented cases are considerably increased compared to main analysis, leading to a higher overall effectiveness and efficiency of the booster vaccination as compared to the main analysis.

*Supplementary Figure 7:* ***Expected number of IMD cases by serogroup in each vaccination scenario 1-5 over 10 years, 2020-2029, sensitivity A3 (constant incidence without booster).***

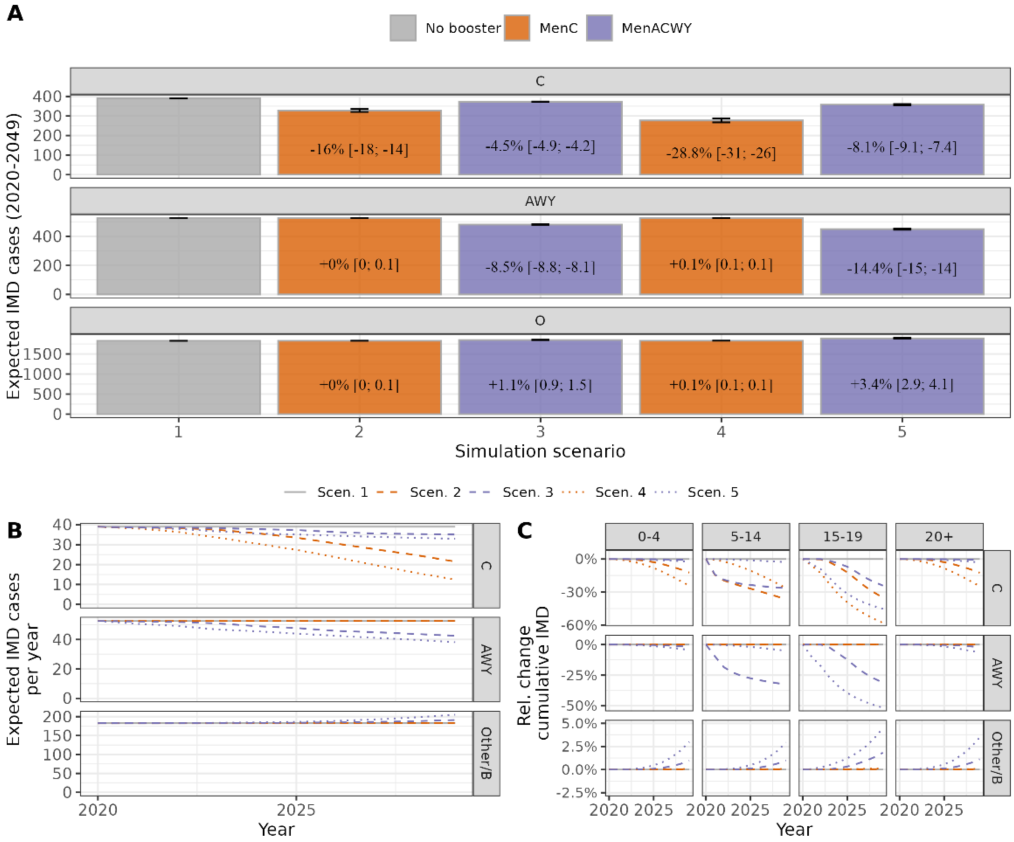


Expected case numbers and absolute numbers of prevented cases are considerably increased for MenC and Other/B (no downward trend) and decreased for MenAWY (no upward trend) compared to main analysis. This leads to an increased effectiveness and efficiency of the MenC booster vaccination as well as a reduced effectiveness and efficiency of the MenACWY booster vaccination as compared to the main analysis.

*S3 B: Model fit*

The choice of model fit used during simulation was less impactful for the MenACWY booster, while effectiveness of the MenC booster was decreased due overall fewer extrapolated MenC cases and also fewer prevented MenC cases.

*Supplementary Figure 8:* ***Expected number of IMD cases by serogroup in each vaccination scenario 1-5 over 10 years, 2020-2029, sensitivity B (alternative fit).***

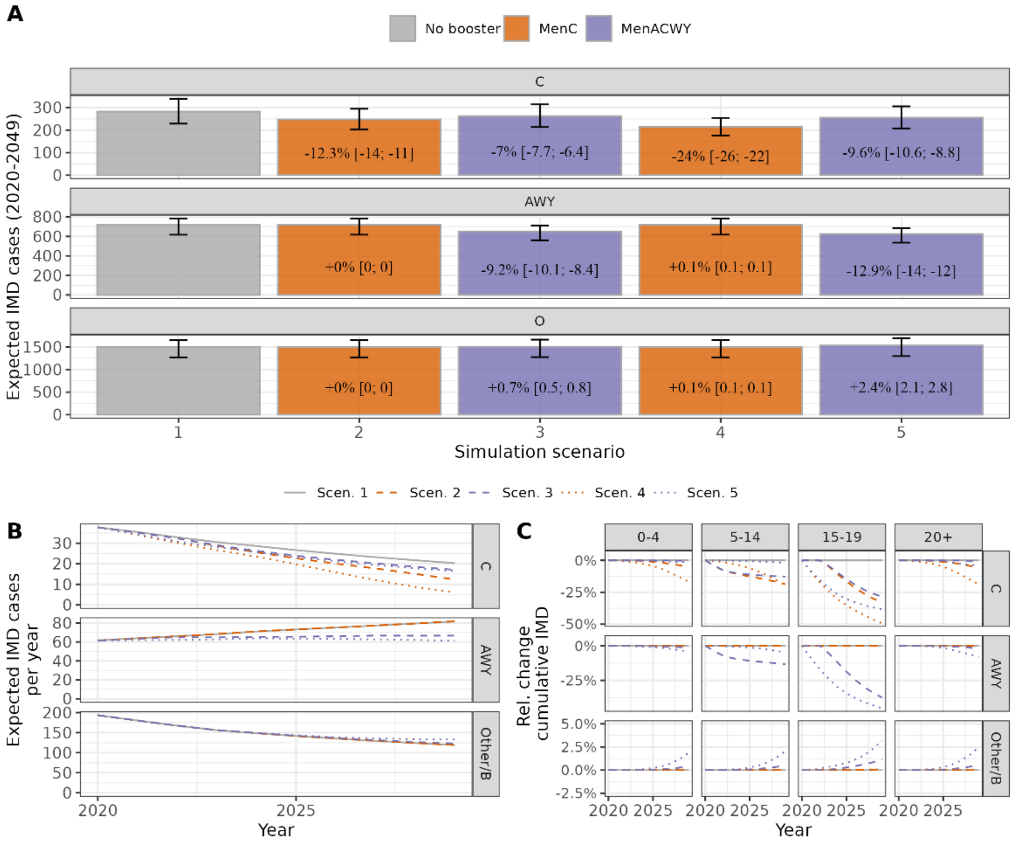

*S3 C: MenACWY protection against carriage*

The assumed degree of protection against carriage of the MenACWY vaccine had direct consequences for the MenACWY booster simulations: assuming no protection against carriage led to a reduction of expected prevented IMD cases by 5%, while assuming an 80% VE against carriage increased prevented IMD cases by 11% compared to the main analysis (with 16% VE against carriage) for the booster vaccination in life year 13 (Figure 5). While these aggregated changes were do not appear very large, serogroup-specific changes were quite substantial: the higher the protection against carriage, the larger the expected reduction in expected MenC and MenAWY IMD cases from the MenACWY vaccine. However, also serotype-replacement became larger and led to additional expected IMD cases in the Other/B serogroup (SuppFig. 11). For the vaccination program in life year 16, these replacement effects played an even larger role and efficiency did not consistently increase with a higher assumption of protection against carriage. The serotype replacement at high protection against carrier (e.g. 80%) was so large that the overall effect in terms of prevented IMD cases was smaller than for the lower assumed protection of 16% in the main analysis (SuppFig. 11).

*Supplementary Figure 9:* ***Expected number of IMD cases by serogroup in each vaccination scenario 1-5 over 10 years, 2020-2029, sensitivity C1 (ACWY VE against carriage: 0%).***

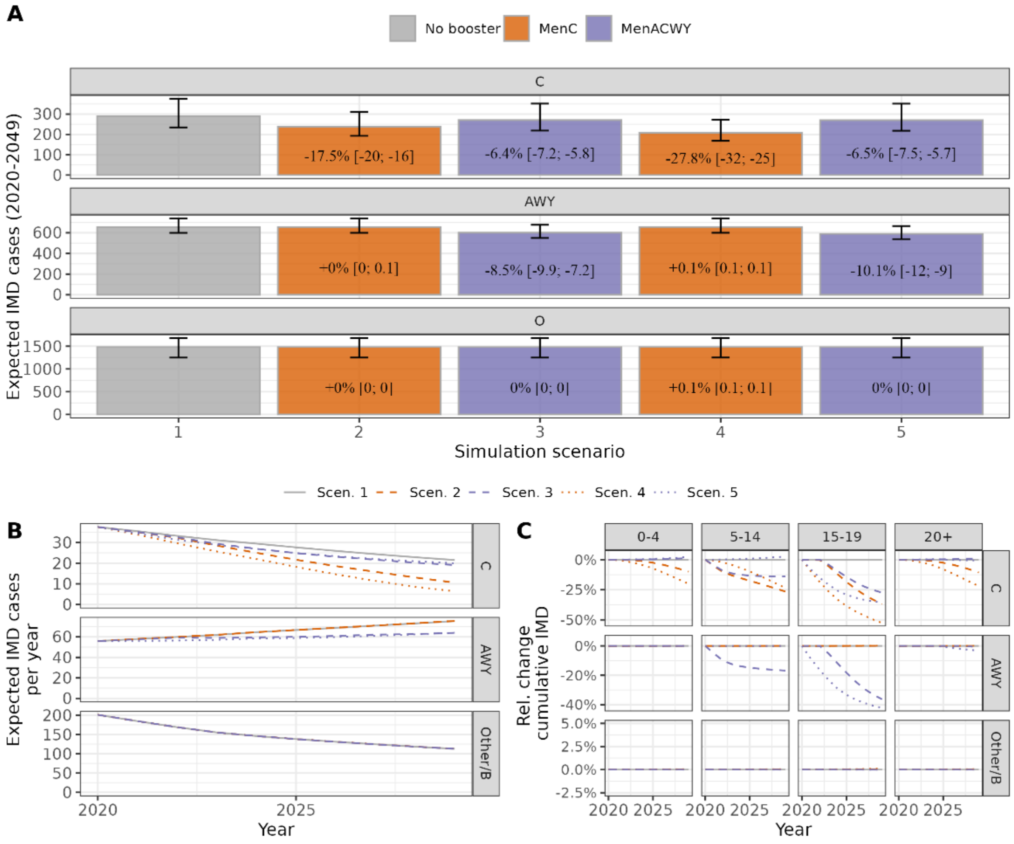


The reduction in expected MenC/AWY IMD cases was reduced compared to the main analysis. For the MenACWY vaccine, there was no strain replacement (increase in expected Other/B cases) without protection against carriage.

*Supplementary Figure 10: **Expected number of IMD cases by serogroup in each vaccination scenario 1-5 over 10 years, 2020-2029, sensitivity C2 (ACWY VE against carriage: 80%).***

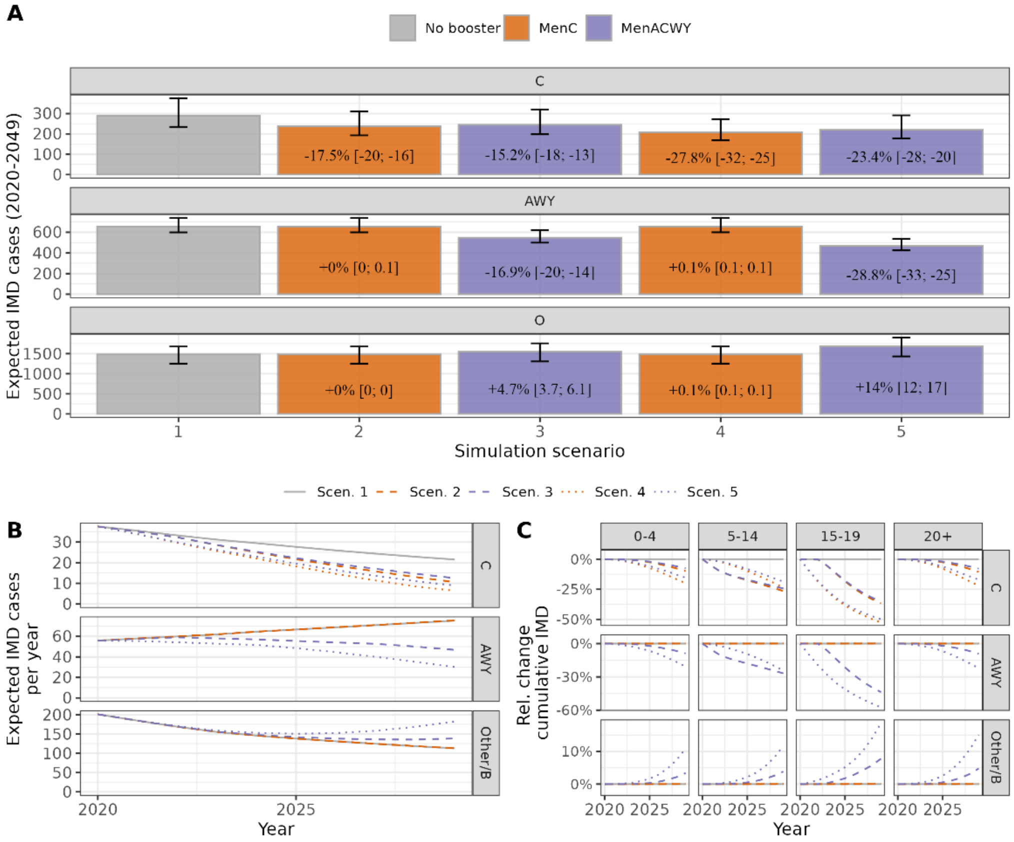


The reduction in expected MenC/AWY IMD cases was increased compared to the main analysis. For the MenACWY vaccine, strain replacement (increase in expected Other/B cases) was more pronounced with increased protection against carriage, in particular for the booster vaccination in life year 16 (simulation scenario 5).

*Supplementary Figure 11: Expected number of prevented IMD cases by serogroup and overall in the 10-year simulation period depending on the assumed protection against carriage for the ACWY booster, booster in life year 13 (scenario 3, panel A) and life year 16 (scenario 5, panel B).*

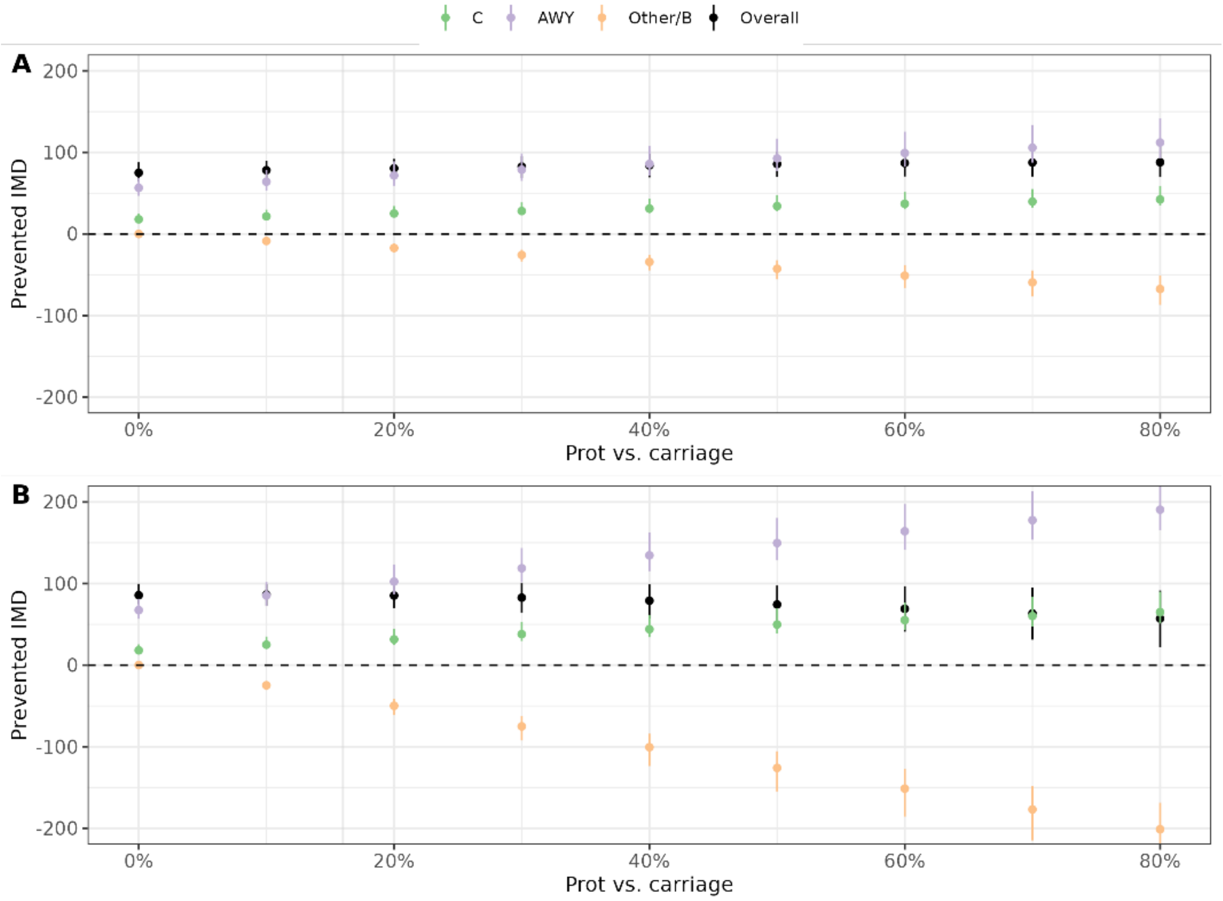

*S3 D: Further parameters/assumptions*

Assuming a shorter duration of protection for the adolescent booster (mean protection 4 years instead of 10, D1, Supplementary Figure 12), lead to reduced effectiveness and efficiency of the adolescent booster (rather consistently for the MenC and MenACWY booster, Fig. 5, SuppFig. 4), while increasing the assumed duration of carriage from 6 to 12 month (D2, Supplementary Figure 13) resulted in minor differences only.

We also investigated the effect of varying the case-carrier ratio. In the first sensitivity analysis (D3), we used the same data for specifying the CCR as compared to the main analysis but different modeling assumptions and extrapolation outside the data supporting leading mainly to a larger case-carrier ratio among infants (age 0-2). In the second analysis (D4) we used data on meningococcal carriage by age from a systematic review that reported higher carriage prevalence in age groups 0–15 years and 50+ in compared to the German data used in the main analysis, which in turn led to substantially lower CCRs, especially at a young age (Supplementary Text 1 – Case carrier ratio, *Supplementary Figure ST2).* Results of the simulations based on the alternative case-carrier ratios showed that the corresponding assumptions in our analyses can have a substantial influence on the results. In analysis D3, the simulated effectiveness of MenC and MenACWY booster vaccination was increased by 3% and 6% for the MenC and MenACWY booster in life year 13, and 2% and 13% for the booster in life year 17, respectively (Figure 5, Supplementary Figure 4, Supplementary Figure 14). In analysis D4, the consequences for MenC and MenACWY vaccination were less consistent. In particular, the MenACWY vaccination showed a marked change: with 120 (booster in life year 13, 95%-UI: 108, 146) and 160 (life year 16, 95%-UI: 145, 191) instead of 77 (life year 13, 95%-UI: 67, 91) and 83 (live year 16, 95%-UI: 71, 100) prevented IMD cases over the 10-year simulation period, the effectiveness of the ACWY booster was increased by 56% or 92%. These additional cases averted resulted from a higher overall extrapolated number of expected (AWY) IMD cases in the simulation period, consequently more MenACWY IMD cases averted, and in comparison, a relatively smaller number of additional expected Other/B IMD cases (i.e., less serotype replacement in sensitivity analysis D4 compared to the main analysis, Supplementary Figure 15). When calibrating the model using the alternative CCR in analysis D4, carriage prevalence of MenAWY was smaller than carriage prevalence of the Other/B serogroup at the end of the calibration period and beginning of the simulation of the different vaccination strategies. In the main analysis, MenAWY carriage prevalence was much larger compared to the carriage prevalence of serogroup Other/B (Supplementary Figure 16). This explains a larger potential for serotype replacement due to the MenACWY booster vaccine in the main analysis as compared to sensitivity analysis D4.

*Supplementary Figure 12:* ***Expected number of IMD cases by serogroup in each vaccination scenario 1-5 over 10 years, 2020-2029, sensitivity D1 (mean duration booster protection 4y).***

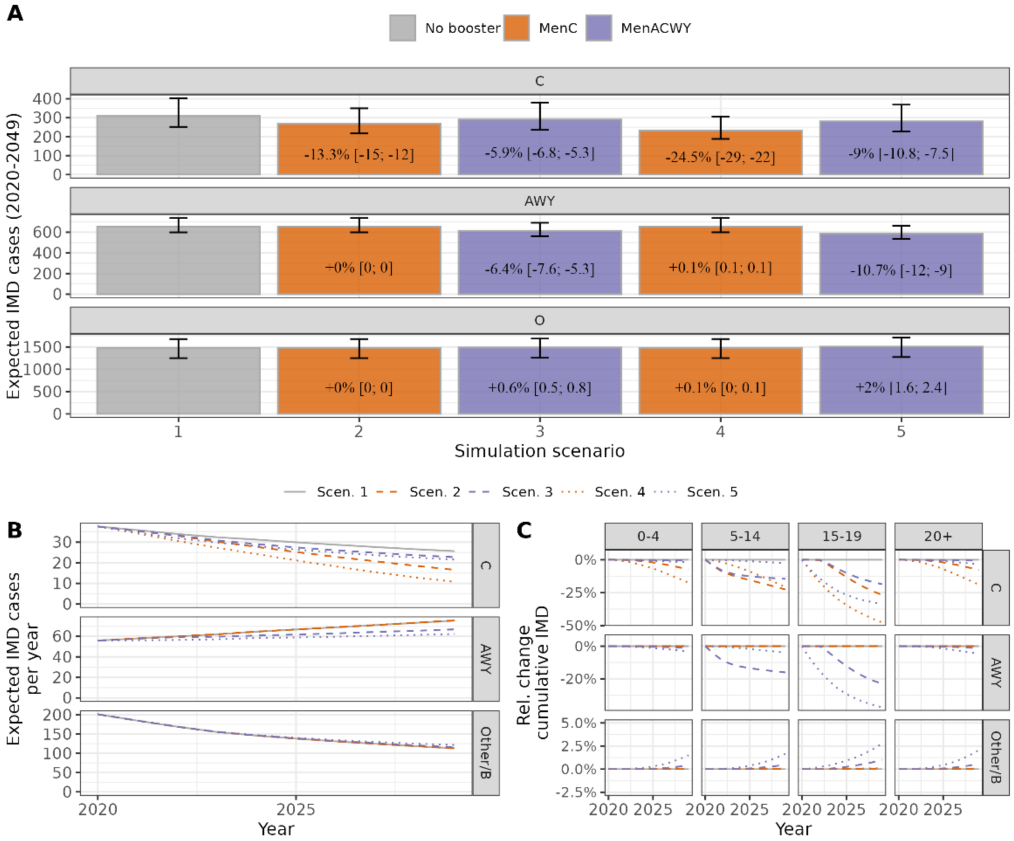


*Supplementary Figure 13:* ***Expected number of IMD cases by serogroup in each vaccination scenario 1-5 over 10 years, 2020-2029, sensitivity D2 (mean carriage duration: 1y).***

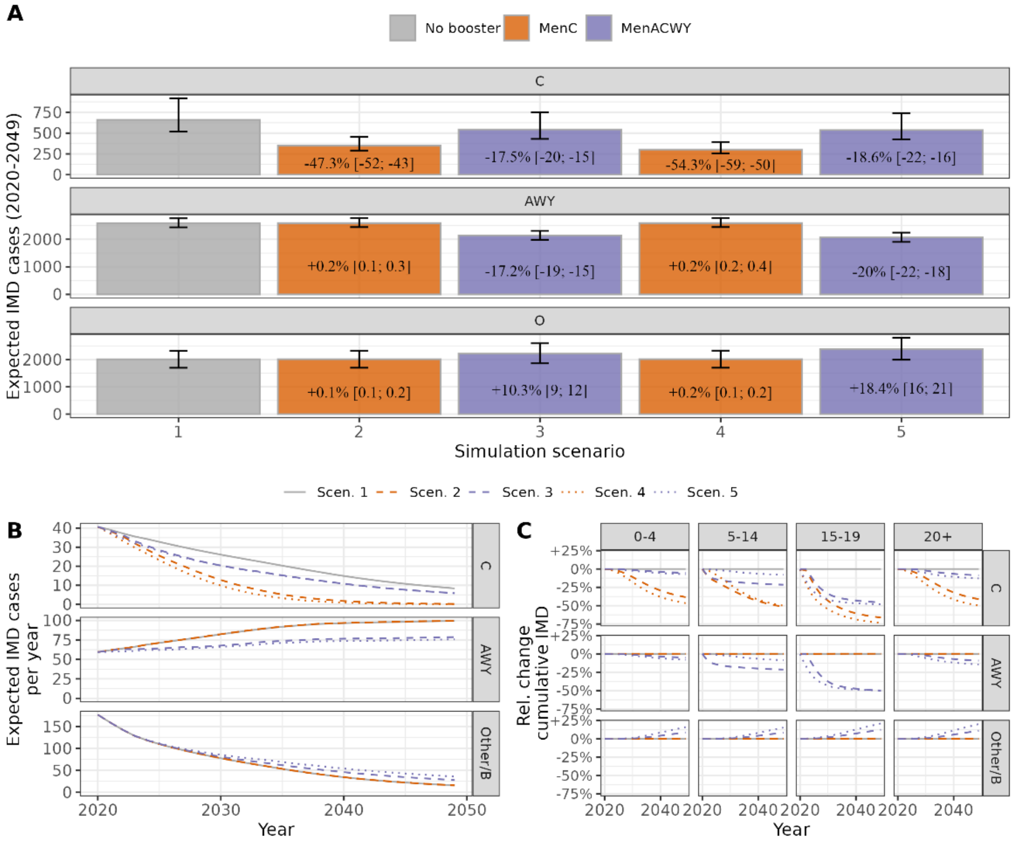

*Supplementary Figure 14:* ***Expected number of IMD cases by serogroup in each vaccination scenario 1-5 over 10 years, 2020-2029, sensitivity D3 (Alternative CCR 1).***

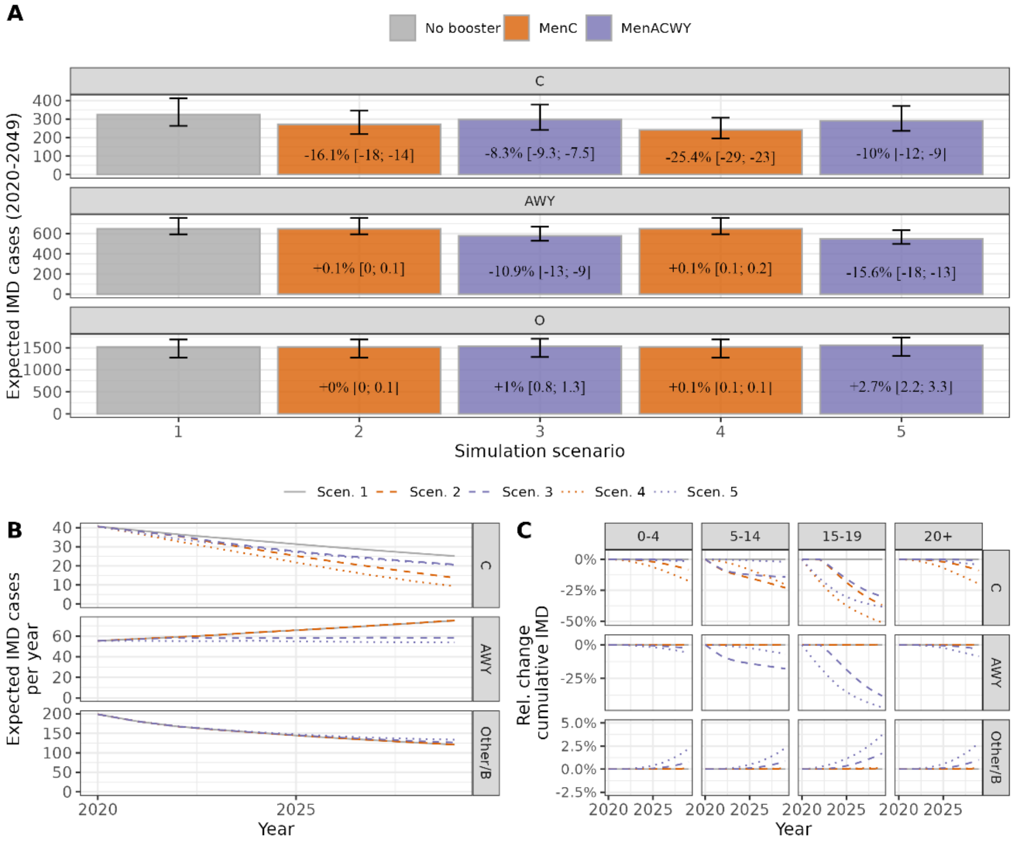


*Supplementary Figure 15:* ***Expected number of IMD cases by serogroup in each vaccination scenario 1-5 over 10 years, 2020-2029, sensitivity D4 (Alternative CCR 2).***

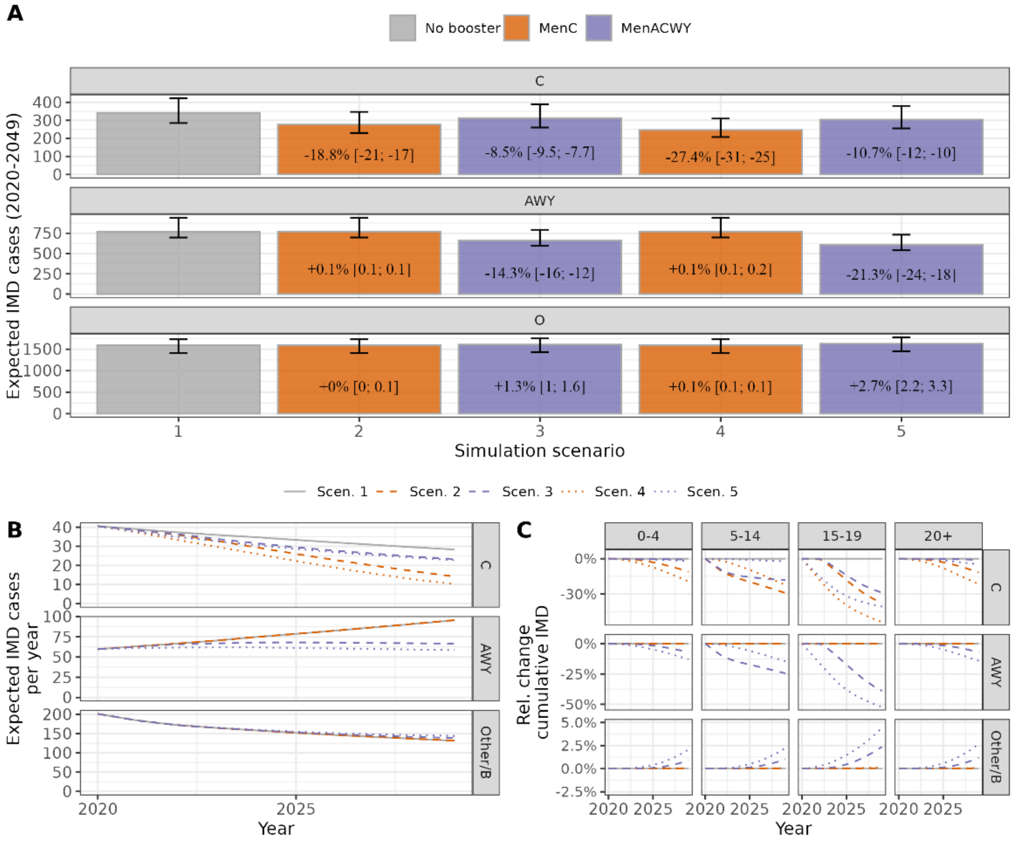

*Supplementary Figure 16: Age-specific meningococcal carriage prevalence by serogroup (rows) in different years of the calibration period (columns) in models calibrated based on different assumptions on the CCR (main analysis, sensitivity analysis D3 (CCR1), and D4 (CCR2), color-coded). Note the different y-axis scales per row.*

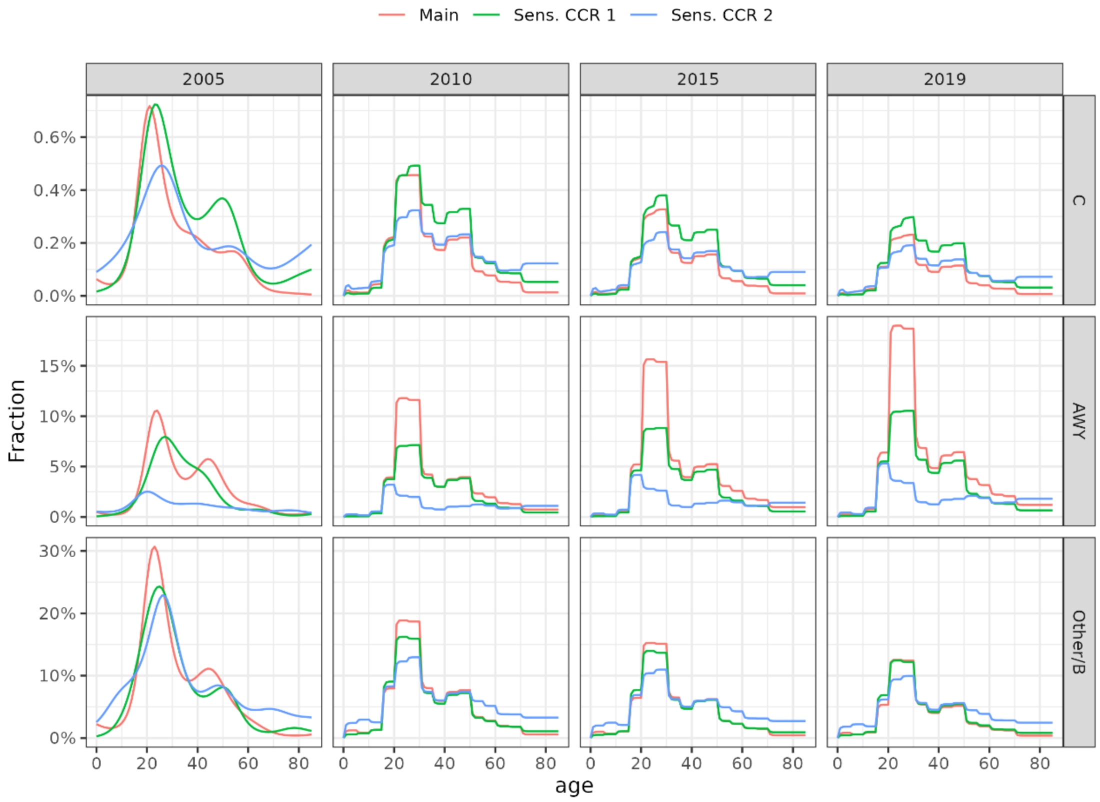

*S3 E: Booster uptake*

Reducing the booster uptake to 60% among all individuals with primary immunization (sensitivity E) led to a reduction of prevented IMD cases by 30-40% (Figure 5, SuppFig. 4, SuppFig. 17). However, since the number of vaccine doses was also reduced by 40%, the efficiency of the vaccination programs remained relatively constant or even increased slightly.

*Supplementary Figure 17:* ***Expected number of IMD cases by serogroup in each vaccination scenario 1-5 over 10 years, 2020-2029, sensitivity E.***

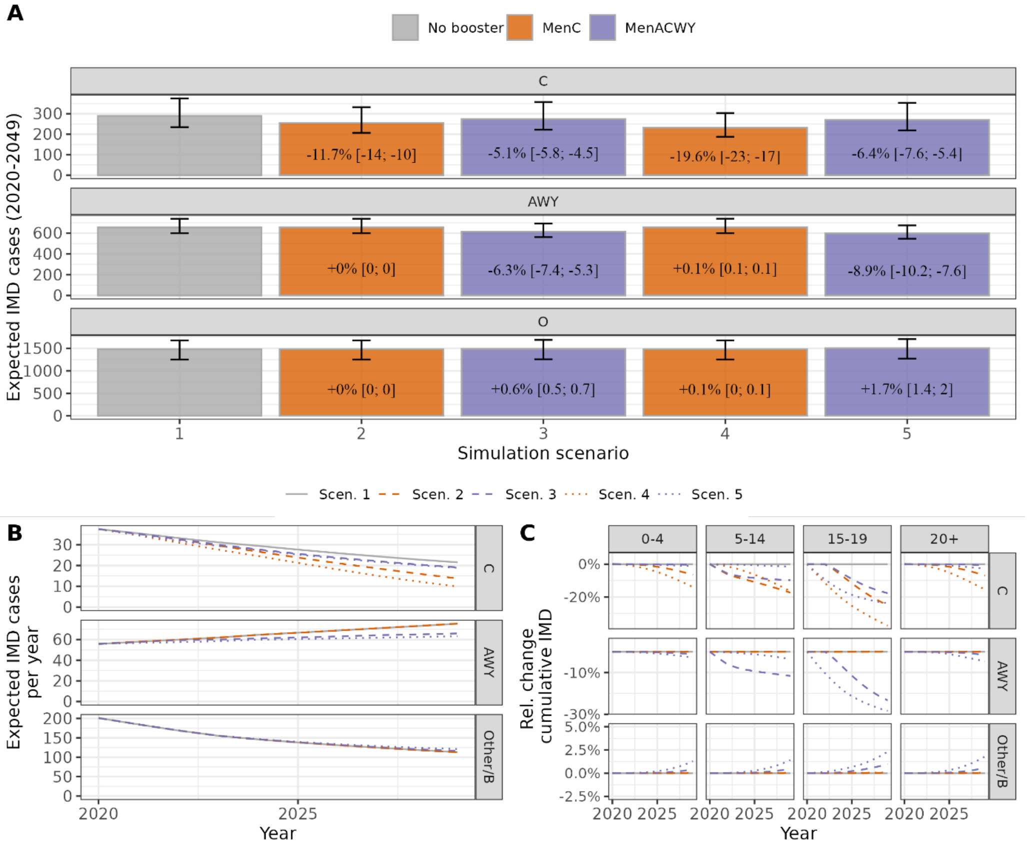

#### Extended simulation period of 30 years

*Supplementary Table 5: Model results of the effectiveness and efficiency of introducing the adolescent booster vaccination program in Germany over 30 years excluding other/B serogroup (as compared to no-booster program, scenario 1). In case of a negative effect (more expected cumulative case numbers compared to the no-booster simulation, scenario 1) the NNV is undefined and marked as "-".*

| Scenario | Outcome | Prevented total number of outcomes (95%-UI) | | | Numbers needed to vaccinate to prevent one outcome (95%-UI) | | |
|---|---|---|---|---|---|---|---|
| | | Mean | Lower 95%-UI | Upper 95%-UI | Mean | Lower 95%-UI | Upper 95%-UI |
| 2 | IMD cases | 240 | 160 | 380 | 81,000 | 52,000 | 120,000 |
| | Sequalae or death | 100 | 70 | 160 | 190,000 | 120,000 | 280,000 |
| | Death | 29 | 20 | 46 | 670,000 | 420,000 | 980,000 |
| 3 | IMD cases | 14 | -130 | 180 | 1,400,000 | 110,000 | - |
| | Sequalae or death | 7 | -52 | 72 | 2,700,000 | 270,000 | - |
| | Death | 3 | -9 | 17 | 5,900,000 | 1,200,000 | - |
| 4 | IMD cases | 290 | 200 | 440 | 69,000 | 45,000 | 100,000 |
| | Sequalae or death | 120 | 84 | 190 | 160,000 | 100,000 | 230,000 |
| | Death | 35 | 24 | 54 | 560,000 | 360,000 | 820,000 |
| 5 | IMD cases | -330 | -510 | -86 | - | - | - |
| | Sequalae or death | -130 | -200 | -33 | - | - | - |
| | Death | -24 | -38 | -3 | - | - | - |

*Supplementary Figure 18:* ***Expected number of IMD cases by serogroup in each vaccination scenario 1-5 over 30 years, 2020-2029****. Cumulative total number of cases and relative change compared to scenario 1 (panel A), annual number of cases by serogroup (panel B) and relative change in the cumulative number of cases by sero- and age-group (panel C). Vertical dotted line represents the end of the 10-year simulation period.*

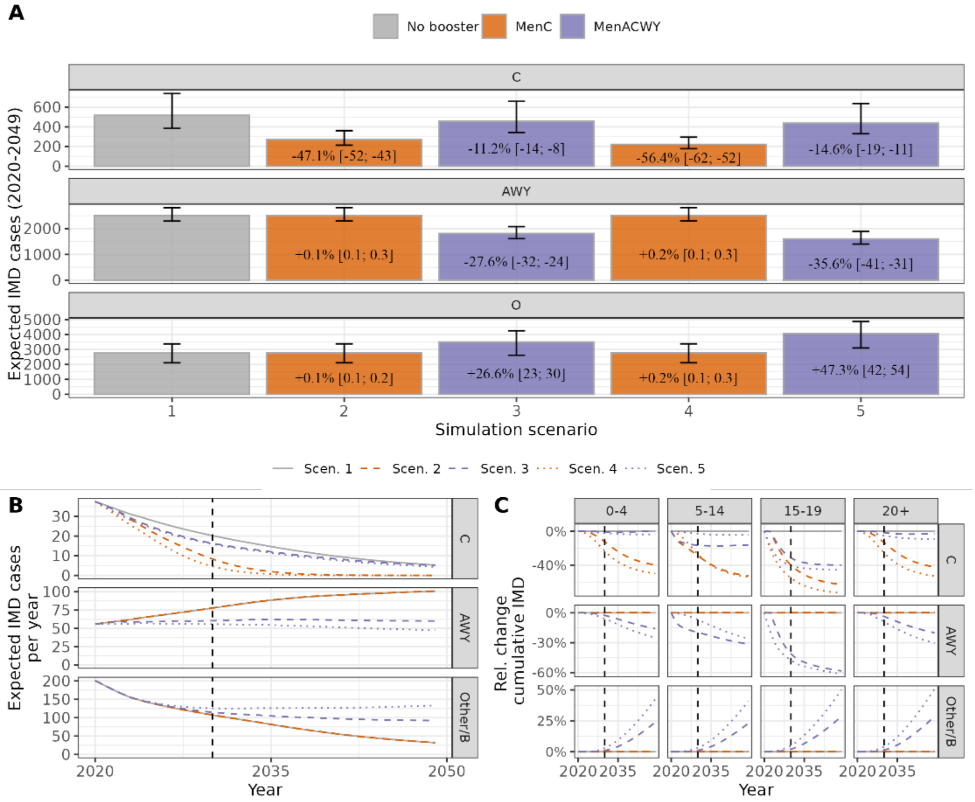

*Supplementary Table 6: Serogroup-specific changes in expected outcomes (prevented cases) for the different adolescent booster scenarios compared to simulation scenario 1 (no booster) over 30 years.*

| | | IMD | | | Sequelae or Death | | | Death | | |
|---|---|---|---|---|---|---|---|---|---|---|
| Scen. | Sero. | Prev. | 2.5%-UI | 97.5%-UI | Prev. | 2.5%-UI | 97.5%-UI | Prev. | 2.5%-UI | 97.5%-UI |
| | C | 246,5 | 168,1 | 386,6 | 105,2 | 71,7 | 165,3 | 29,7 | 20,2 | 47,2 |
| 2 | AWY | -3,8 | -6,4 | -2,5 | -1,5 | -2,6 | -1,0 | -0,3 | -0,6 | -0,2 |
| | Other/B | -3,0 | -5,1 | -1,6 | -1,2 | -2,0 | -0,6 | -0,3 | -0,4 | -0,1 |
| | C | 60,0 | 37,7 | 85,4 | 25,2 | 15,8 | 36,0 | 6,7 | 4,1 | 9,6 |
| 3 | AWY | 692,1 | 599,8 | 805,6 | 280,2 | 243,0 | 326,1 | 60,2 | 52,4 | 70,0 |
| | Other/B | -737,9 | -922,3 | -526,3 | -298,4 | -372,5 | -213,0 | -63,7 | -79,0 | -45,8 |
| | C | 295,0 | 202,0 | 452,0 | 126,0 | 86,2 | 193,5 | 35,8 | 24,4 | 55,5 |
| 4 | AWY | -4,7 | -7,6 | -3,1 | -1,9 | -3,1 | -1,3 | -0,4 | -0,7 | -0,3 |
| | Other/B | -4,2 | -6,9 | -2,3 | -1,7 | -2,8 | -0,9 | -0,4 | -0,6 | -0,2 |
| | C | 79,3 | 46,7 | 116,0 | 33,7 | 19,8 | 49,3 | 9,3 | 5,5 | 13,7 |
| 5 | AWY | 900,1 | 778,0 | 1020,8 | 365,5 | 316,0 | 414,3 | 80,1 | 70,2 | 90,6 |
| | Other/B | -1309,4 | -1532,9 | -972,5 | -529,5 | -619,7 | -393,7 | -113,1 | -131,4 | -84,6 |

*Supplementary Figure 19: Expected number of prevented IMD cases based on a MenACWY booster in life year 13 (scenario 3) and 16 (scenario 5) compared to the no-booster scenario 1 over a 30-year simulation period by serogroup and overall (color-coded). Show are results depending on different assumptions regarding the protection against ACWY carriage (x-axis) and for different assumptions regarding the case carrier ratio (main analysis, and defined as in sensitivity analysis D3 and D4). Assumed protection against carriage in the main analysis was 16% among individuals with vaccine protection.*

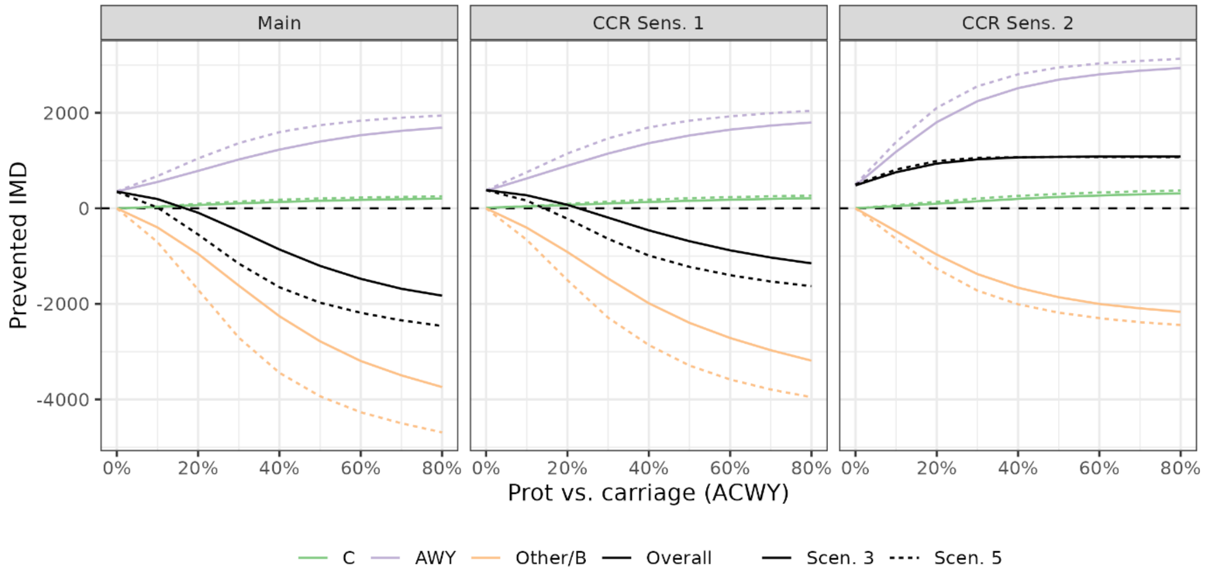
